# Prevalence of Enteric Hyperoxaluria in Crohn’s Disease, Cystic Fibrosis, and After Bariatric Surgery: A Systematic Review

**DOI:** 10.64898/2026.10.07.26364847

**Authors:** Xiaomin Xing, Huiqiong He, Haifeng Liu, Qingshan Li

## Abstract

**Background:** Enteric hyperoxaluria (EH) causes calcium oxalate kidney stones, oxalate nephropathy, and chronic kidney disease (CKD), and can progress to end-stage kidney disease. It occurs in Crohn’s disease (CD), cystic fibrosis (CF), chronic pancreatitis, celiac disease, short bowel syndrome, and after bariatric surgery. Its prevalence in these populations has not been synthesized.

**Methods:** PROSPERO CRD420261364424. PubMed and Embase were searched to 24 August 2026 for studies reporting 24-hour urinary oxalate. Prevalence was pooled using the Freeman–Tukey double arcsine transformation with a DerSimonian–Laird random-effects model; unweighted crude prevalence is also reported. Because heterogeneity was high, the narrative synthesis stratified by testing condition is the primary result. Prespecified subgroups addressed oxalate load before collection, diagnostic threshold, ileal resection, age, and urine collection format.

**Results:** Forty-seven studies were included. In CD (17 studies, 471 patients; 204 with EH), crude prevalence was 43.3% and pooled prevalence 49.3% (95% CI 38.9–59.8; I² = 95.3%). With an oxalate load, prevalence was 64.7% (pooled 74.0%, 55.9–88.6) versus 38.6% without (pooled 39.1%, 28.7–50.0). Among CD patients, EH was more frequent with ileal resection (48.8% vs 31.7%; OR 1.62, 0.91–2.89) and with steatorrhea (62.5% vs 29.3%; OR 9.88, 3.81–25.61). In CF (7 studies, 348 patients; 179 with EH), pooled prevalence was 49.8% (40.5–59.1; I² = 91.9%). After bariatric surgery (15 studies, 849 patients; 474 with EH), pooled prevalence was 54.7% (45.4–63.9; I² = 96.7%).

**Conclusions:** Roughly half of these patients have EH, but the proportion depends heavily on whether an oxalate load precedes urine collection (39% unprovoked vs 65%–74% provoked); because most studies do not state this condition, published prevalence estimates are not directly comparable. Ileal resection and steatorrhea identify those who should be screened.

**Key Points:**

1. Enteric hyperoxaluria was present in about half of patients with Crohn’s disease, cystic fibrosis, or prior bariatric surgery.
2. Measured prevalence nearly doubled when an oxalate load preceded urine collection (74% vs 39%), so published estimates are not comparable.
3. Ileal resection and steatorrhea identified patients at higher risk and can guide who should be screened.

## Introduction

Enteric hyperoxaluria (EH) is a secondary metabolic disorder whose central mechanism is the failure of intraluminal calcium sequestration caused by fat malabsorption, with a consequent increase in colonic oxalate absorption ^1,2^. Unabsorbed fatty acids and bile salts chelate calcium in the lumen, so dietary oxalate can no longer precipitate as poorly soluble calcium oxalate and be excreted in the feces, raising the concentration of soluble oxalate ^3,4^; they also damage the colonic epithelium and increase permeability, promoting oxalate absorption ^5,6^. Absorbed oxalate enters the portal circulation, is freely filtered by the glomerulus, and is excreted in the urine, where it binds calcium to form a highly supersaturated calcium oxalate species — the strongest driver of calcium oxalate stone formation ^6,7^. Beyond malabsorptive bariatric surgery, Crohn’s disease (CD), cystic fibrosis (CF), chronic pancreatitis (CP), celiac disease (CeD), and short bowel syndrome (SBS) cause EH by the same route: each either damages the small intestinal mucosa directly or causes exocrine pancreatic insufficiency, and all converge on impaired fat absorption ^1^.

The consequences are not confined to urinary oxalate. As in primary hyperoxaluria, crystals deposit in the renal parenchyma and can lead to oxalate nephropathy, CKD, and ESKD ^1,8^. Among 814 patients with EH followed for a mean of 5 years, 60% to 90% developed kidney stones within 1 to 5 years and 5.7% progressed to ESKD ^9^, and among 108 cases of secondary oxalate nephropathy 55% required dialysis and none recovered renal function ^10^. Such outcomes occur only in patients who were tested for EH.

This bears directly on how many patients should be expected. Measuring urinary oxalate requires a 24-hour collection, is rarely undertaken outside specialist centres, and produces no symptom that would prompt testing, so kidney stones and CKD cannot serve as a proxy for EH: they are the downstream consequence in the minority of patients who were both tested and followed long enough to develop them. The only published attempt to count this population illustrates the difficulty. Using a state-transition Markov model on Truven claims data — that is, from diagnostic codes rather than from urinary oxalate — Tasian et al.^11^ estimated that approximately 250,000 people in the United States with an enteric disorder or a history of enteric surgery also had kidney stones or CKD. That figure describes patients who had already developed kidney complications, and its distribution across diseases reflects the size of each disease population rather than the frequency of EH within it: RYGB accounted for approximately 64%, inflammatory bowel disease 20%, CeD 8%, CP 4%, and SBS the remainder.

Because testing is rarely performed, how common EH is within each of these disease groups has not been established. We therefore systematically searched PubMed/Medline and Embase for studies that measured urinary oxalate in patients with CD, CF, CP, CeD, or SBS, or after malabsorptive bariatric surgery, and synthesized their findings.

## Methods

### Search strategy and eligibility criteria

This review followed the PRISMA (Preferred Reporting Items for Systematic Reviews and Meta-Analyses) 2020 statement, and the protocol was prospectively registered with PROSPERO (CRD420261364424). We searched PubMed and Embase from inception to 24 August 2026 using terms that included “hyperoxaluria” and the name of each target disease. Title and abstract screening was performed by the principal investigator (PI) and, in parallel, by an independently run large-language-model-assisted workflow (DeepSeek V4-Flash, 21 August 2026); agreement on definite include/exclude decisions was κ = 0.86, and records judged uncertain by either reviewer proceeded to full-text assessment. Full-text assessment and data extraction were performed by the PI, with every value checked against the original full text and screenshots retained.

Eligibility criteria were: patients with confirmed CD, CF, CP, CeD, or SBS, or who had undergone malabsorptive bariatric surgery (predominantly Roux-en-Y gastric bypass, RYGB); at least one 24-hour urinary oxalate measurement per patient, or an oxalate-to-creatinine ratio convertible to a 24-hour rate; a reported prevalence of hyperoxaluria or counts from which it could be calculated; and a sample size of at least 5. We excluded primary hyperoxaluria, animal studies, reviews, conference abstracts, and case reports, and treated SBS as a secondary, descriptive population because too few studies were available.

### Data extraction

For each study we extracted first author, year, country, disease and subgroup (ileal resection, steatorrhea, age), sample size, number with EH, urinary oxalate concentration and units, assay method, diagnostic threshold, and the dietary conditions before collection. Mixed-etiology studies that could not be disaggregated were tabulated separately and excluded from pooling. All values were checked against the original full text; unreported values were recorded as NR and not imputed.

### Statistical analysis

Pooled proportions used the Freeman–Tukey double arcsine transformation ^12^: for c events among n participants, T = asin√(c/(n+1)) + asin√((c+1)/(n+1)) with SE = √(1/(4(n+1))). Estimates used a DerSimonian–Laird random-effects model ^13^ with weights 1/(SE² + τ²), back-transformed as p = [sin(T/2)]²; heterogeneity was assessed with Cochran’s Q, I², and τ². We also report the unweighted crude prevalence (ΣEH/Σn), which conveys the distribution of the raw data and avoids over-weighting small studies.

Prespecified subgroups addressed oxalate load before testing, diagnostic threshold, ileal resection, age, and urine collection format. Pooled estimates were calculated for subgroups with k ≥ 3 and crude prevalence only for k ≤ 2; mixed-etiology subgroups were not pooled, and robustness was examined by leave-one-out analysis. The PROSPERO protocol (item 4.8.2) specifies that when I² ≥ 75%, pooling may not be appropriate and narrative synthesis should be the primary result. All three pooled estimates had I² > 90%, so in accordance with that prespecified rule we present the narrative synthesis stratified by pre-test diet and threshold as the primary result and the random-effects estimates as exploratory summaries, not as prevalences applicable to any single setting. Analysis used Python 3.11 (NumPy).

## Results

### Study selection

The two databases yielded 1,185 records (PubMed 365; Embase 820); after removal of 312 duplicates, 873 remained. Title and abstract screening excluded 682, leaving 191 for full-text assessment, of which 97 were excluded (64 full text unavailable, 21 conference abstracts, 10 not meeting eligibility criteria, 2 content mismatch) and 94 read. Excluding a further 44 studies with no EH data (32), stone outcomes only (9), or other reasons (3) gave 49 EH prevalence data points from 47 independent studies. Thirty-nine provided single-disease pooled estimates (CD 17, CF 7, bariatric surgery 15) and 8 (CP 1, exocrine pancreatic insufficiency 1, CeD 3, SBS 3) were reported descriptively because too few studies were available (Figure 1).

**Figure 1.**
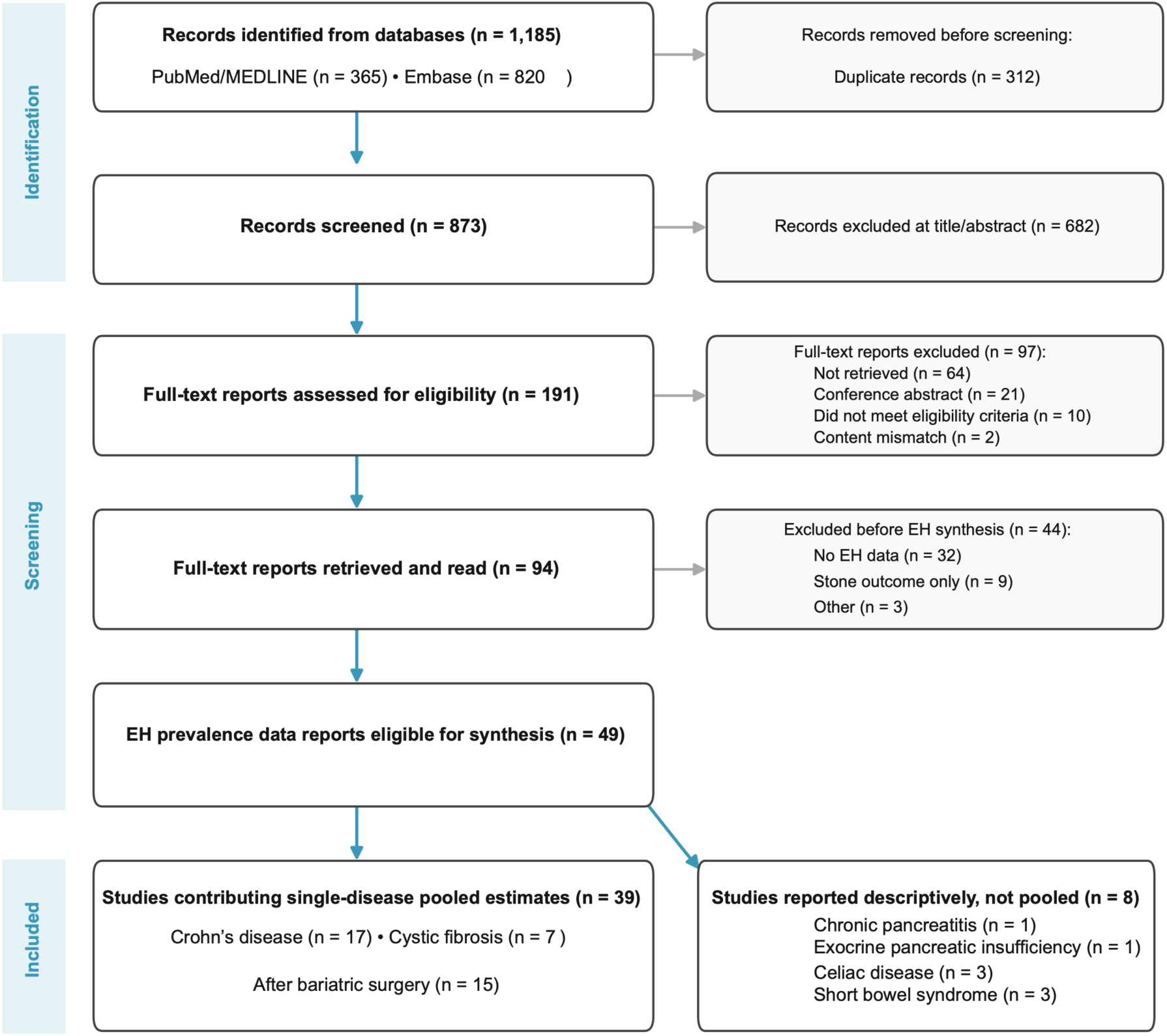
Literature screening flowchart following the PRISMA 2020 protocol. The search was performed on 24 August 2026. EH, enteric hyperoxaluria. Data points (n = 49) exceed independent studies (n = 47) because two studies contributed to two disease groups.

### Crohn’s disease

CD most often involves the terminal ileum and impairs fat absorption, while weight loss makes overlap with the bariatric population unlikely. Seventeen studies measured urinary oxalate in patients with CD (Figure 2; Table 1). Among 471 patients who completed 24-hour collection, 204 met the diagnostic threshold used in their study, a crude prevalence of 43.3% and a pooled prevalence of 49.3% (95% CI 38.9–59.8; I² = 95.3%). Crude prevalence was 50.7% (34/67) in the six US studies and 42.1% (170/404) elsewhere, but the groups were unbalanced and this was not tested.

**Figure 2.**
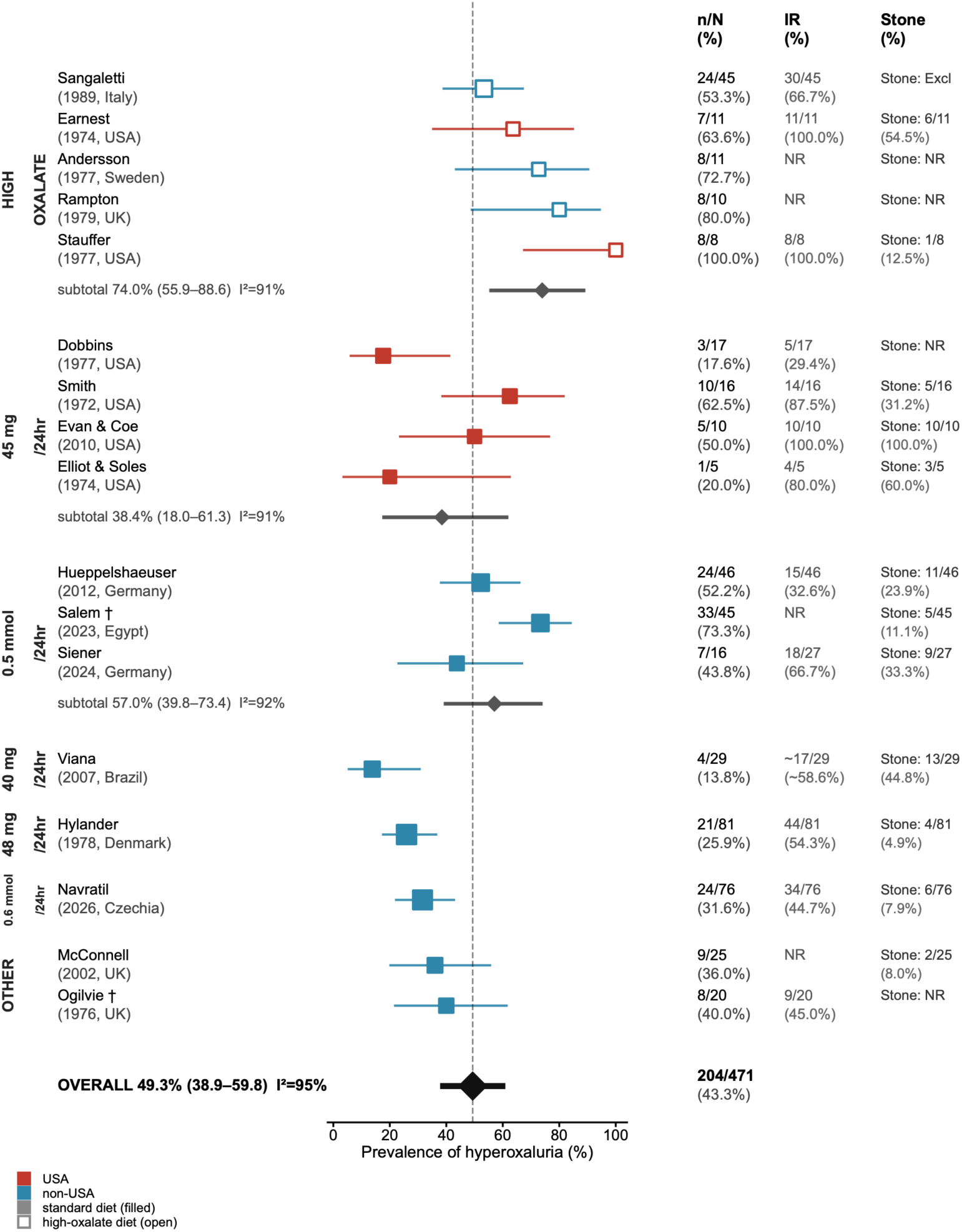
Prevalence of enteric hyperoxaluria in patients with Crohn’s disease (k = 17). Individual study proportions with 95% confidence intervals, grouped by whether an oxalate load was given before urine collection. Marker area is proportional to study sample size. Filled markers indicate testing on a habitual or standardized diet and open markers indicate testing after a high-oxalate load. Red markers denote studies from the United States and blue markers studies from other countries. Columns show the number with hyperoxaluria over the group total (n/N), the proportion with hyperoxaluria (%), the proportion with a history of ileal resection (IR), and the number with kidney stones (Stone; Excl = not assessed in that study, NR = not reported). Subtotal and overall estimates were obtained using the Freeman–Tukey double arcsine transformation with a DerSimonian–Laird random-effects model.

**Table 1.**
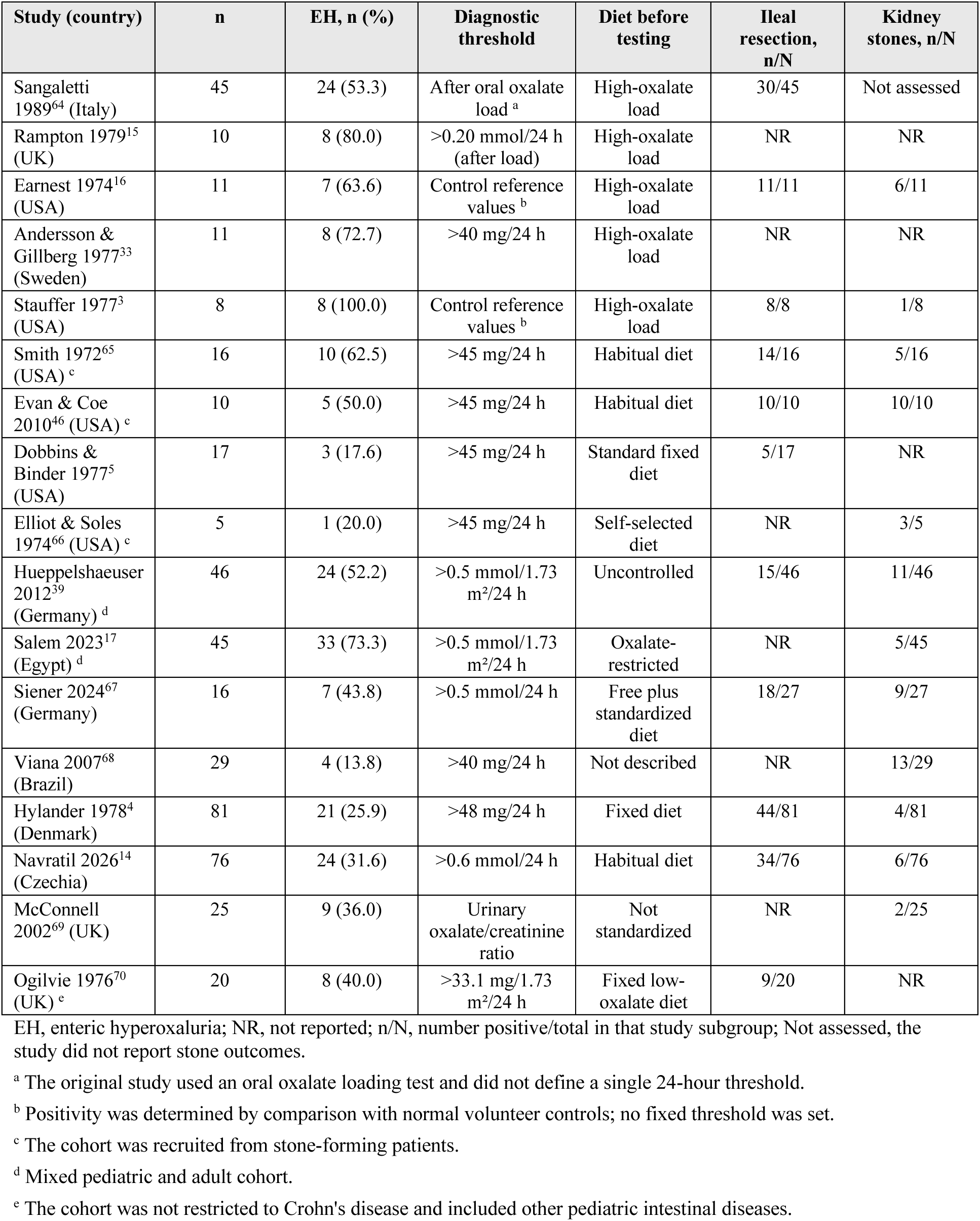
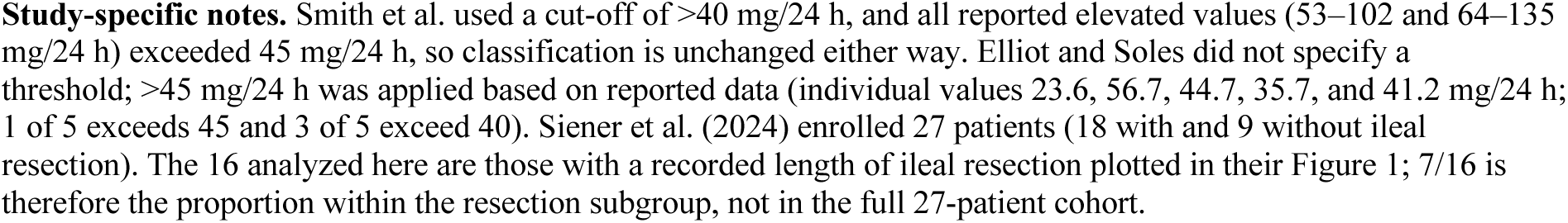
Characteristics of Crohn’s disease studies and prevalence of EH (k = 17)

#### Testing condition and threshold

Five studies gave an oxalate load before collection (85 patients) and twelve tested on a habitual or standardized diet (386 patients; Figure 2; Table 1); crude prevalence was 64.7% (55/85) versus 38.6% (149/386), and pooled estimates 74.0% (95% CI 55.9–88.6; I² = 91.2%) versus 39.1% (28.7–50.0; I² = 94.6%). The 26- to 35-point gap makes the oxalate load the strongest effect modifier in the available evidence, and means the figure of 49.3% for all 17 studies is a mixture of provoked and unprovoked data. Thresholds also differed (Table 2). Under unprovoked conditions, the four US studies using >45 mg/24 h gave 39.6% (19/48; pooled 38.4%, 18.0–61.3; I² = 90.8%) and the two German adult studies at an equivalent threshold 50.0% (31/62); together, 45.5% (50/110) and a pooled 42.1% (29.1–55.6; I² = 87.2%), an interval overlapping that of all 17 studies, so varying threshold and diet explains only part of the heterogeneity. Under unprovoked conditions the proportion with CD and EH therefore falls broadly between 38% and 50%, rising to 65%–74% with an oxalate load. Two studies using higher thresholds reported correspondingly lower proportions, 25.9% (21/81) and 31.6% (24/76) (Figure 2). Salem, at 0.5 mmol/1.73 m², reported 73.3% (33/45) against 50.0% (31/62) in the German adult studies at the same threshold, so that stratum is heavily influenced by one pediatric cohort (Table 2; Figure 2).

**Table 2.** Stratified pooled prevalence of EH in Crohn’s disease by testing condition and diagnostic threshold.

| Group | k | Crude prevalence ($\Sigma EH/\Sigma n$) | Pooled prevalence (95% CI) | I<sup>2</sup> | Studies included |
| --- | --- | --- | --- | --- | --- |
| Oxalate load before testing (provoked) | 5 | 64.7% (55/85) | 74.0% (55.9–88.6) | 91.2% | Sangaletti, Rampton, Earnest, Andersson, Stauffer |
| Habitual diet (unprovoked) | 12 | 38.6% (149/386) | 39.1% (28.7–50.0) | 94.6% | Remaining 12 studies |
| >45 mg/24 h | 4 | 39.6% (19/48) | 38.4% (18.0–61.3) | 90.8% | Smith, Evan & Coe, Dobbins & Binder, Elliot & Soles |
| >0.5 mmol/24 h (or equivalent body-surface-area-corrected value)<sup>b</sup> | 3 | 59.8% (64/107) | 57.0% (39.8–73.4) | 92.1% | Hueppelshaeuser, Salem, Siener |
| >40 mg/24 h | 1 | 13.8% (4/29) | Not pooled (k < 3) | — | Viana |
| >48 mg/24 h | 1 | 25.9% (21/81) | Not pooled (k < 3) | — | Hylander |
| >0.6 mmol/24 h | 1 | 31.6% (24/76) | Not pooled (k < 3) | — | Navratil |
| Oxalate/creatinine ratio or other basis<sup>c</sup> | 2 | 37.8% (17/45) | Not pooled (k < 3) | — | McConnell, Ogilvie |
| Unprovoked with equivalent threshold (45 mg/24 h ∪ 0.5 mmol/24 h)<sup>a</sup> | 6 | 45.5% (50/110) | 42.1% (29.1–55.6) | 87.2% | Smith, Evan & Coe, Dobbins & Binder, Elliot & Soles, Hueppelshaeuser, Siener |
| All studies | 17 | 43.3% (204/471) | 49.3% (38.9–59.8) | 95.3% | — |

<sup>a</sup> Habitual or standardized diet only, threshold >45 mg/24 h or its 0.5 mmol/24 h equivalent. <sup>b</sup> Groups by threshold value: Hueppelshaeuser 2012 and Salem 2023 used >0.5 mmol/1.73 m<sup>2</sup>/24 h (body-surface-area-corrected); Siener 2024a used >0.5 mmol/24 h. <sup>c</sup> McConnell 2002 (oxalate/creatinine ratio) and Ogilvie 1976 (>33.1 mg/1.73 m<sup>2</sup>/24 h, not expressible in the mmol rows above). "Not pooled (k < 3)" indicates that no stratum-specific pooled estimate was computed because fewer than three studies were available (see Methods); those studies were nevertheless included in the "All studies" estimate. The threshold strata cover only the 12 studies tested on a habitual or standardized diet.

#### Ileal resection

In the studied CD patients, among the eight studies with resection stratification (Table 3), the resection subgroup gave 48.8% (81/166; pooled 58.5%, 42.9–73.3; I² = 94.0%) and the non-resected subgroup 31.7% (32/101; pooled 29.7%, 20.4–39.9; I² = 75.6%), a difference of 17.1 points crude and 28.8 pooled, with urinary oxalate correlating with the length resected under fixed dietary conditions ^4^. Navratil et al.^14^ provide a counterexample: 26.5% (9/34) with ileocolic resection versus 35.7% (15/42) without, and 29.0% (9/31) of healthy controls also met the threshold (Figure 2).

**Table 3.** Prevalence of EH in ileal resection and non-resection subgroups.

| Study (country) | Resection group EH, n/N | Non-resection group EH, n/N |
| --- | --- | --- |
| Smith 1972<sup>65</sup> (USA) | 10/14 | 0/2 |
| Earnest 1974<sup>16</sup> (USA) | 7/11 | — (no such subgroup) |
| Stauffer 1977<sup>3</sup> (USA) | 8/8 | — (no such subgroup) |
| Hylander 1978<sup>4</sup> (Denmark) | 16/44 | 2/11 |
| Sangaletti 1989<sup>64</sup> (Italy) | 17/30 | 7/15 |
| Evan & Coe 2010<sup>46</sup> (USA) | 5/10 | — (no such subgroup) |
| Hueppelshaeuser 2012<sup>39</sup> (Germany) | 9/15 | 8/31 |
| Navratil 2026<sup>14</sup> (Czechia) | 9/34 | 15/42 |
| **Total** | 81/166 (48.8%) | 32/101 (31.7%) |
| **Pooled prevalence (95% CI)** | 58.5% (42.9–73.3) | 29.7% (20.4–39.9) |
| **I<sup>2</sup>** | 94.0% | 75.6% |
| **τ<sup>2</sup>** | 0.189 | 0.041 |

EH, enteric hyperoxaluria; —, subgroups not present in that study. Pooled values were estimated using the Freeman–Tukey double arcsine transformation with a DerSimonian–Laird random-effects model; crude prevalence is the unweighted ΣEH/Σn. The remaining nine studies were not included because of lacking information.

Steatorrhea. Among the five CD studies reporting both measures (Rampton 1979,^15^, Earnest 1974,^16^, Dobbins and Binder 1977,^5^, Hylander 1978,^4^, and Salem 2023,^17^; 164 patients), steatorrhea was present in 62.5% (45/72) of patients with EH and 29.3% (27/92) of those without, a difference of 33.2 points (Mantel-Haenszel OR 9.88, 95% CI 3.81–25.61). Definitions of steatorrhea varied (fecal fat cut-offs from >6 g/d to >50 g/72 h), so this is directional only and was not pooled.

### Cystic fibrosis

CF is an autosomal recessive disorder caused by mutations in *CFTR* (cystic fibrosis transmembrane conductance regulator), a chloride channel, with an incidence in Caucasian populations long estimated at 1 in 2,500 live births ^18^. CFTR dysfunction causes exocrine pancreatic insufficiency and fat malabsorption, predisposing to EH ^19,20^; approximately 39,000 people in the United States have CF ^21^.

Seven studies measured urinary oxalate in 348 patients with CF (the 20 heterozygote parents reported by Terribile et al. served as an independent control group and were not included), of whom 179 had hyperoxaluria, a crude prevalence of 51.4%; the random-effects pooled prevalence was 49.8% (95% CI 40.5–59.1; I² = 91.9%) (Figure 3; Table 4). One further report from the same group (Hoppe et al., 1998) was excluded because the later study states that 21 of its 37 patients had been followed since 1993 and were therefore included in previously published work, indicating overlap between the two cohorts. Proportions ranged from 35.0% (Niknahad 14/40) to 69.9% (Andrieux 58/83, Figure 3; Table 4). Terribile et al.20 reported hyperoxaluria in 14 of 29 patients with homozygous CF (48.3%), 3 of 20 heterozygotes (15%), and 1 of 30 controls (3.3%), suggesting a dose-related association with loss of CFTR function; Andrieux et al.^19^ found it associated with pancreatic insufficiency and severe genotypes.^22^

**Figure 3.**
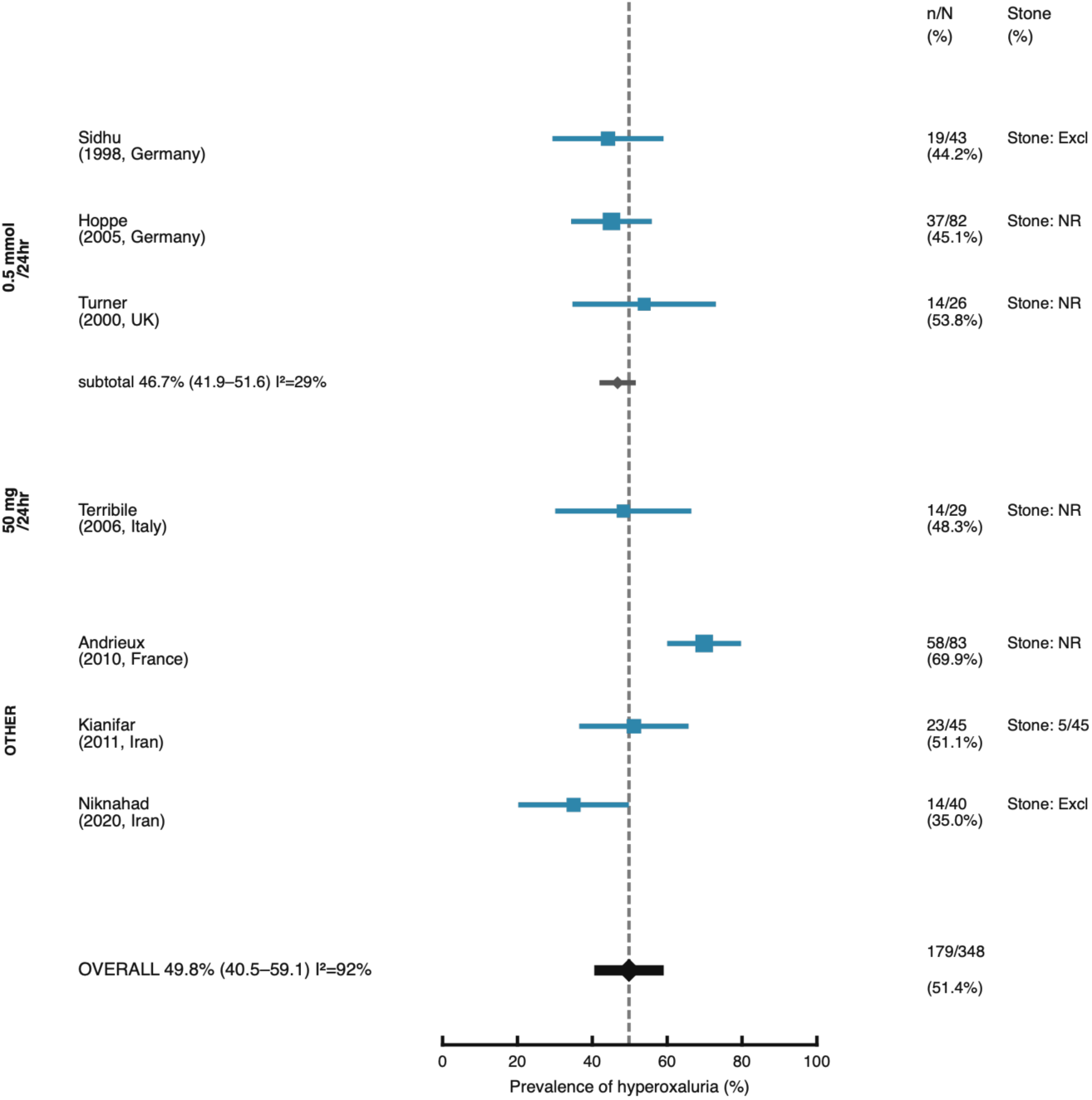
Prevalence of enteric hyperoxaluria in patients with cystic fibrosis (k = 7). Individual study proportions with 95% confidence intervals, grouped by diagnostic threshold. Marker area is proportional to study sample size. Columns show the number with hyperoxaluria over the group total (n/N), the proportion with hyperoxaluria (%), and the number with kidney stones (Stone; Excl = not assessed in that study, NR = not reported). All studies measured urinary oxalate on a habitual or oxalate-restricted diet; none used an oxalate load. Subtotal and overall estimates were obtained using the Freeman–Tukey double arcsine transformation with a DerSimonian–Laird random-effects model.

**Table 4.** Characteristics of cystic fibrosis studies and prevalence of EH (k = 7)

| Study (country) | n | EH, n (%) | Age | Diagnostic threshold | Kidney stones, n/N |
| --- | --- | --- | --- | --- | --- |
| Sidhu 1998<sup>37</sup> (Germany) | 43 | 19 (44.2) | 3–39 y | >0.5 mmol/24 h | NR |
| Hoppe 2005<sup>71</sup> (Germany) | 82 | 37 (45.1) | 5–37 y | >0.5 mmol/1.73 m<sup>2</sup>/24 h for 12 mo | 6/37 |
| Turner 2000<sup>72</sup> (UK) | 26 | 14 (53.8) | 5–15.9 y | >0.5 mmol/24 h | 0/26 |
| Terribile 2006<sup>73</sup> (Italy) | 29 | 14 (48.3) | 28.4 ± 7.1 y | Not specified (approximately 50 mg/24 h) | 6/29 |
| Andrieux 2010<sup>19</sup> (France) | 83 | 58 (69.9) | 3.9–13.0 y | Urinary oxalate/creatinine ratio | 2/83 |
| Kianifar 2011<sup>74</sup> (Iran) | 45 | 23 (51.1) | Mean 47.1 months | Not specified | 0/45 |
| Niknahad 2020<sup>60</sup> (Iran) | 40 | 14 (35.0) | 4.34 ± 3.38 y | Composite criteria | 1/40 |
| **Total** | **348** | **179 (51.4)** | — | — | — |
| **Pooled prevalence (95% CI)** | — | **49.8% (40.5–59.1)** | — | — | — |
| **I<sup>2</sup>** | — | **91.9%** | — | — | — |

EH, enteric hyperoxaluria; NR, not reported. The denominator for Hoppe 2005 is the 82 patients with CF under care at that centre, of whom 37 were persistently hyperoxaluric (urinary oxalate >0.5 mmol/1.73 m<sup>2</sup>/24 h for the 12 months before study entry) and were studied in detail; hyperoxaluria is defined by that 12-month criterion rather than by a single 24-hour measurement, so the estimate is the more conservative of the two possible readings of that study.

### Bariatric surgery

Restrictive procedures generally do not cause EH ^23,24^, whereas RYGB is the procedure most often associated with it. Approximately 670,000 gastric bypass operations were performed in the United States between 2011 and 2023 ^25^ and including adjacent years the cumulative number of patients who have undergone RYGB may exceed 800,000. Fifteen studies included 849 patients after bariatric surgery, predominantly RYGB, of whom 474 had EH, a crude prevalence of 55.8% and a pooled prevalence of 54.7% (95% CI 45.4–63.9; I² = 96.7%) (Figure 4; Table 5). Proportions ranged from 19.7% (Froeder 12/61) to 87.0% (Nelson 20/23) and were higher with more malabsorptive procedures: Asplin and Coe^26^ reported abnormal values in 92 of 132 patients (69.7%) and Moreland et al.^27^ in 20 of 26 after RYGB (76.9%) against 5 before surgery. Most studies came from the United States (10 studies, 316/488 = 64.8%) and the rest from Germany and Brazil (2 each) and Canada and Hong Kong (1 each), together 158/361 = 43.8%. The diet was uncontrolled and the threshold most often 40 or 45 mg/24 h.

**Figure 4.**
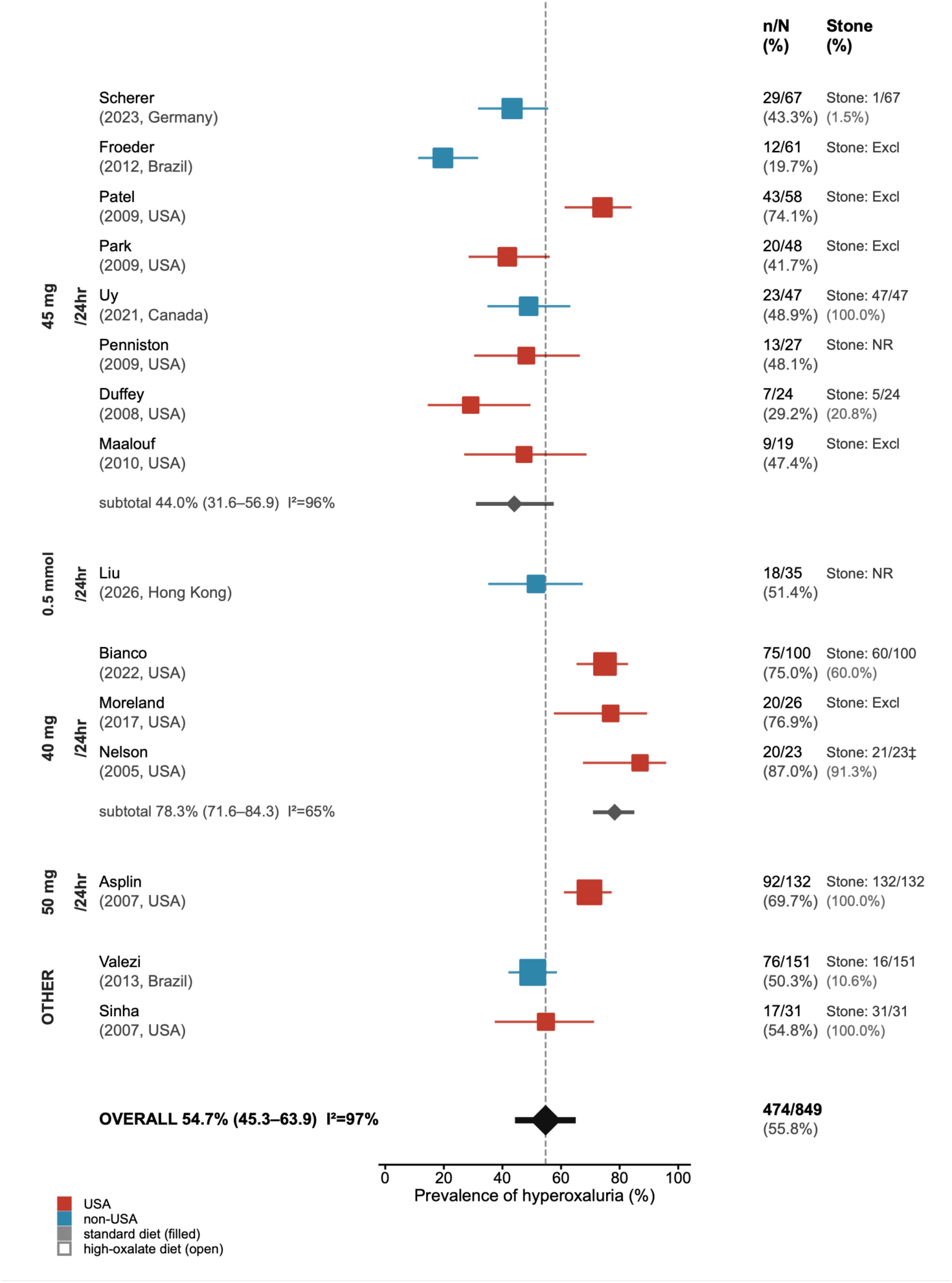
Prevalence of enteric hyperoxaluria after bariatric surgery (k = 15). Individual study proportions with 95% confidence intervals, grouped by procedure type. Marker area is proportional to study sample size. Red markers denote studies from the United States and blue markers studies from other countries. Columns show the number with hyperoxaluria over the group total (n/N), the proportion with hyperoxaluria (%), and the number with kidney stones (Stone; Excl = not assessed in that study, NR = not reported). ‡ the 23 patients were identified by a diagnosis of calcium oxalate nephrolithiasis (n = 21) or oxalate nephropathy (n = 2). Subtotal and overall estimates were obtained using the Freeman–Tukey double arcsine transformation with a DerSimonian–Laird random-effects model.

**Table 5.**
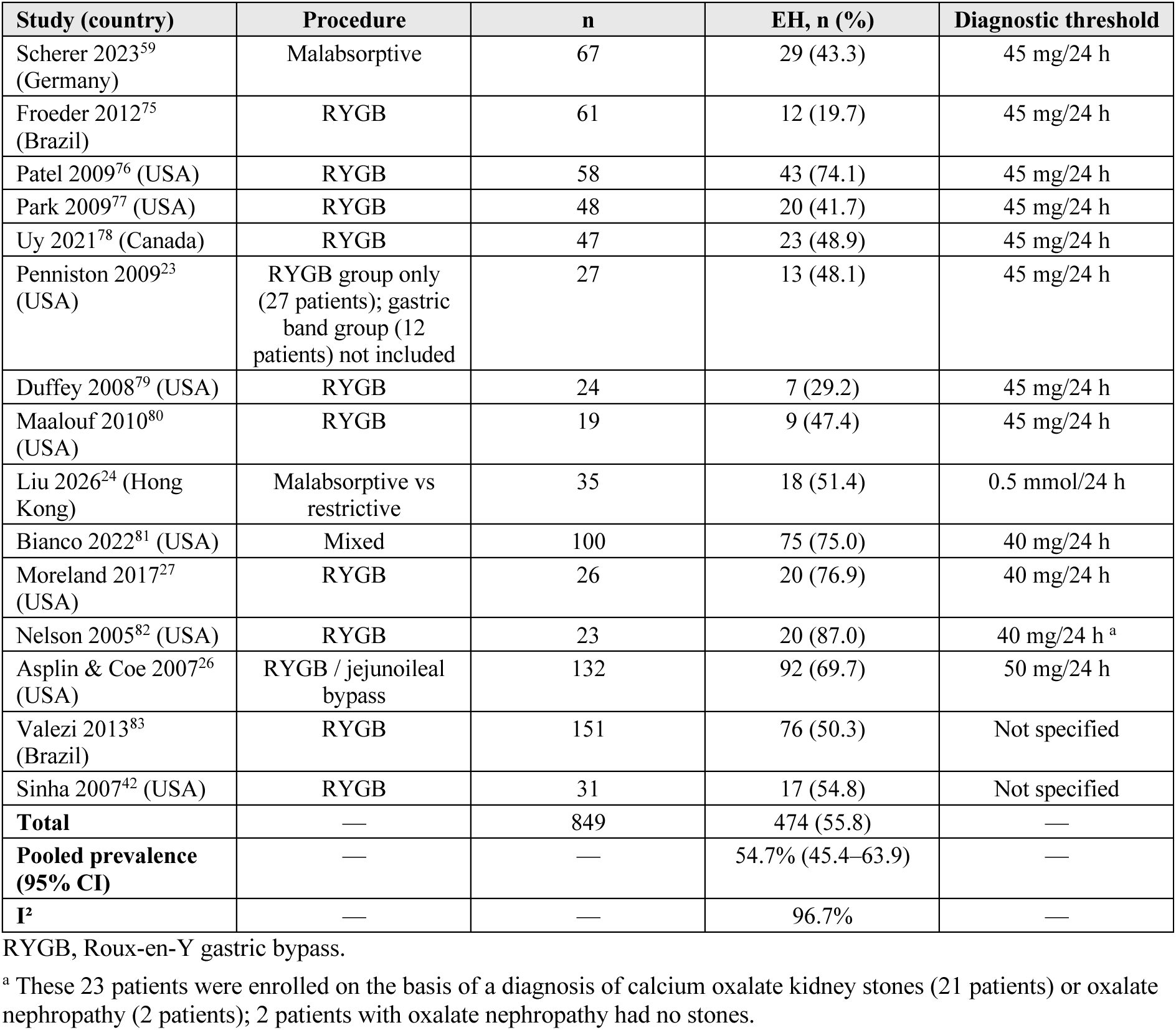
Characteristics of bariatric surgery studies and prevalence of EH (k = 15)

| Study (country) | Procedure | n | EH, n (%) | Diagnostic threshold |
| --- | --- | --- | --- | --- |
| Scherer 2023<sup>59</sup> (Germany) | Malabsorptive | 67 | 29 (43.3) | 45 mg/24 h |
| Froeder 2012<sup>75</sup> (Brazil) | RYGB | 61 | 12 (19.7) | 45 mg/24 h |
| Patel 2009<sup>76</sup> (USA) | RYGB | 58 | 43 (74.1) | 45 mg/24 h |
| Park 2009<sup>77</sup> (USA) | RYGB | 48 | 20 (41.7) | 45 mg/24 h |
| Uy 2021<sup>78</sup> (Canada) | RYGB | 47 | 23 (48.9) | 45 mg/24 h |
| Penniston 2009<sup>23</sup> (USA) | RYGB group only (27 patients); gastric band group (12 patients) not included | 27 | 13 (48.1) | 45 mg/24 h |
| Duffey 2008<sup>79</sup> (USA) | RYGB | 24 | 7 (29.2) | 45 mg/24 h |
| Maalouf 2010<sup>80</sup> (USA) | RYGB | 19 | 9 (47.4) | 45 mg/24 h |
| Liu 2026<sup>24</sup> (Hong Kong) | Malabsorptive vs restrictive | 35 | 18 (51.4) | 0.5 mmol/24 h |
| Bianco 2022<sup>81</sup> (USA) | Mixed | 100 | 75 (75.0) | 40 mg/24 h |
| Moreland 2017<sup>27</sup> (USA) | RYGB | 26 | 20 (76.9) | 40 mg/24 h |
| Nelson 2005<sup>82</sup> (USA) | RYGB | 23 | 20 (87.0) | 40 mg/24 h <sup>a</sup> |
| Asplin & Coe 2007<sup>26</sup> (USA) | RYGB / jejunioileal bypass | 132 | 92 (69.7) | 50 mg/24 h |
| Valezi 2013<sup>83</sup> (Brazil) | RYGB | 151 | 76 (50.3) | Not specified |
| Sinha 2007<sup>42</sup> (USA) | RYGB | 31 | 17 (54.8) | Not specified |
| **Total** | — | 849 | 474 (55.8) | — |
| **Pooled prevalence (95% CI)** | — | — | 54.7% (45.4–63.9) | — |
| **I<sup>2</sup>** | — | — | 96.7% | — |

RYGB, Roux-en-Y gastric bypass.
<sup>a</sup> These 23 patients were enrolled on the basis of a diagnosis of calcium oxalate kidney stones (21 patients) or oxalate nephropathy (2 patients); 2 patients with oxalate nephropathy had no stones.

### Chronic pancreatitis, celiac disease, and short bowel syndrome

For these three diseases, and for exocrine pancreatic insufficiency, too few studies were available for pooling (Table 6).

**Table 6.**
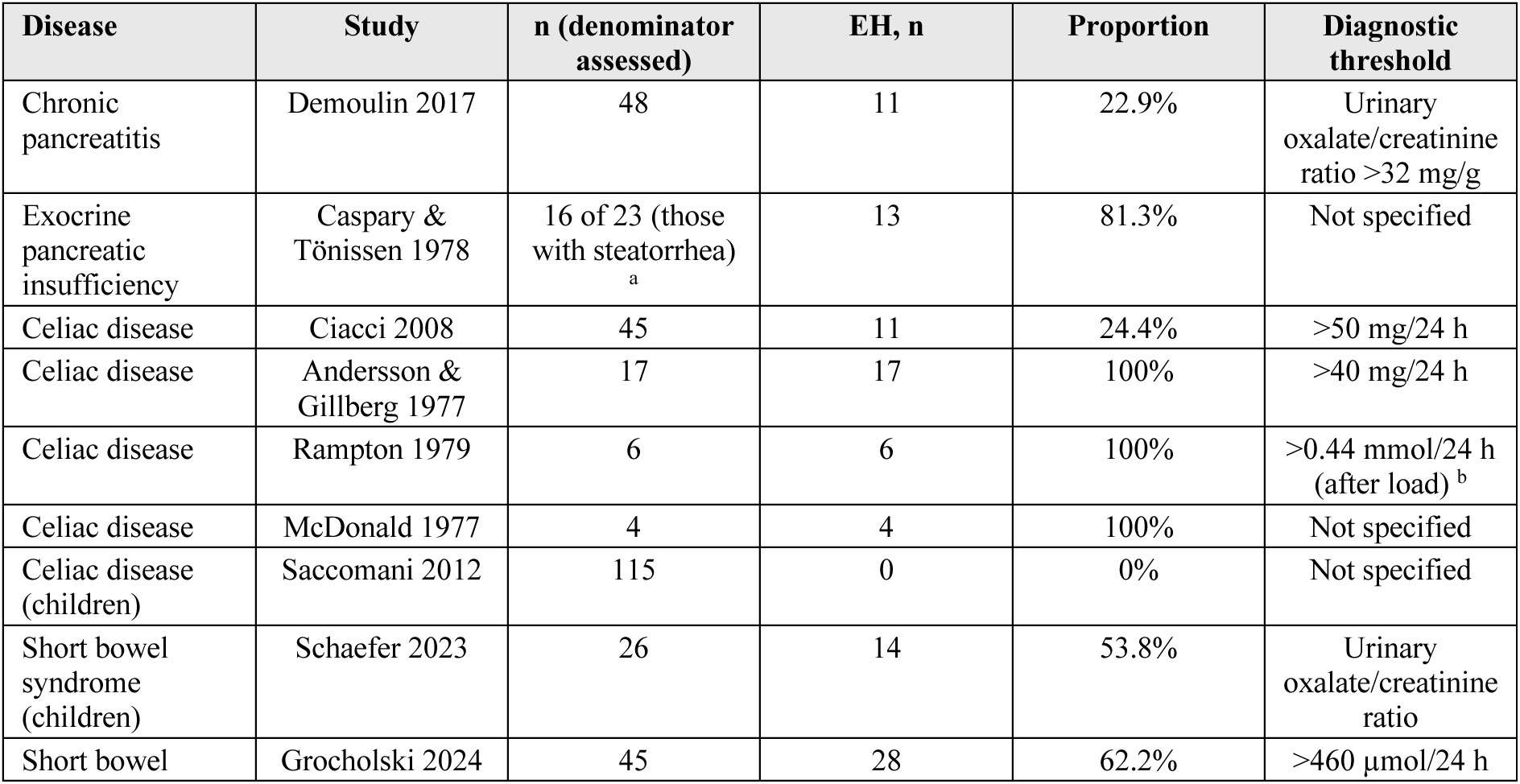
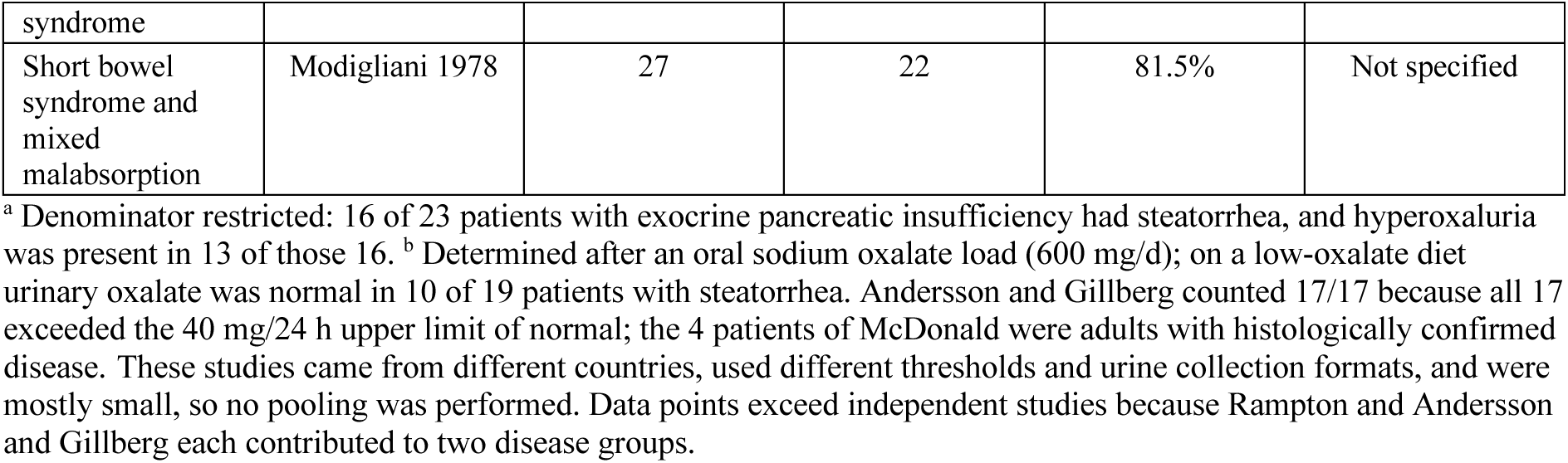
Prevalence of EH in pancreatic disease, celiac disease, and short bowel syndrome.

#### Chronic pancreatitis and exocrine pancreatic insufficiency

Demoulin et al.^28^ found EH in 11 of 48 patients with CP (22.9%), identified with a positive predictive value of 100% and a negative predictive value of 86% by the combination of clinical steatorrhea, a fecal acid steatocrit >31%, and pancreatic atrophy. Caspary and Tönissen^29^ reported hyperoxaluria in 13 of the 16 of 23 patients with exocrine pancreatic insufficiency who had steatorrhea (81.3%), and Cartery et al.^30^ in all 12 patients with CP-related acute oxalate nephropathy, of whom 3 progressed to ESKD.

#### Celiac disease

Ciacci et al.^31^ reported hyperoxaluria (>50 mg/24 h) in 11 of 45 adults with CeD (24.4%); all four patients of McDonald et al.^32^ and all six of Rampton et al.^15^ were hyperoxaluric, and the 17 patients of Andersson and Gillberg^33^ had a mean urinary oxalate of 102 (41–185) mg/24 h, all above the normal limit. By contrast, Saccomani et al.^34^ found none among 115 children with CeD, suggesting that risk differs between adults and children.

#### Short bowel syndrome

Schaefer et al.^35^ reported EH in 14 of 26 children with SBS (53.8%) and Grocholski et al.^36^ in 28 of 45 patients with SBS (62.2%), with lower estimated glomerular filtration rate in the hyperoxaluric group; Modigliani et al.^7^ reported hyperoxaluria in 22 of 27 patients with malabsorption due to ileal resection (81.5%).

## Discussion

### Mechanisms

Oxalate is an end product of human metabolism excreted mainly in the urine, derived from dietary intake and endogenous hepatic production ^1,2^. Dietary oxalate normally binds calcium in the colon to form poorly soluble calcium oxalate that is excreted in the feces, and blood oxalate can also be secreted into the lumen by the duodenal SLC26A6 transporter ^20^; urinary oxalate therefore reflects both intestinal absorption and hepatic production.

In fat malabsorption, unabsorbed fatty acids and bile salts form soaps with calcium, the calcium sequestration effect is weakened, and oxalate is excessively absorbed, mainly in the colon ^3,5^. Intestinal dysbiosis, such as the absence of colonization with *Oxalobacter formigenes*, and impaired intestinal oxalate secretion also contribute ^20,37,38^.

Three findings bear on this mechanism. Giving an oxalate load raised the detected proportion from 39.1% to 74.0% (Table 2), consistent with substrate supply limiting the measured prevalence. Steatorrhea was present in 62.5% of patients with EH versus 29.3% of those without (Mantel-Haenszel OR 9.88, 95% CI 3.81–25.61), consistent with fat malabsorption as the driver. The EH proportion was 17.1 percentage points higher with ileal resection (48.8% vs 31.7%, Table 3), although this contrast did not reach significance (OR 1.62, 95% CI 0.91–2.89), consistent with reduced absorptive surface contributing alongside, rather than independently of, the other mechanisms.

### Kidney outcomes

Calcium oxalate stones are common in these populations and recur readily. In CD, 6 of 11 patients in Earnest et al.^16^ and 11 of 46 in Hueppelshaeuser et al.^39^ had stones, 8 of the latter recurrent; among 332 patients with CF the lifetime cumulative prevalence was 32% ^40^, with higher urinary oxalate in those with stones ^41^; and after RYGB, 21 of 31 patients had calcium oxalate stones, more than 11 recurrent ^42^.

EH can in turn lead to CKD. CKD was present in 91% of patients with SBS and EH ^36^, EH accounted for 60% of 113 biopsy-confirmed cases of secondary oxalate nephropathy, mostly after RYGB ^8^, and 3 of 12 patients with CP-related oxalate nephropathy progressed to ESKD ^30^. In inflammatory bowel disease, a US national cohort showed increased risks of stones (aOR 2.25) and CKD (aOR 1.28) ^43^, a Swedish cohort reported 10-year cumulative incidences of CKD of 11.8% and stones of 5.6% ^44^, and stones increased the risk of CKD 10-fold among patients with CD and CKD ^45^. Whether detecting or treating EH alters this course requires prospective study.

The damage is not confined to stones in the urinary tract: crystals deposit in the medullary collecting ducts and interstitium, causing injury and inflammation that give way to atrophy and fibrosis ^10,46^, and moderate-to-severe interstitial fibrosis was 3.5 times more likely with an enteric than a non-enteric cause ^47^. Because oxalate is excreted almost entirely by the kidney, the process is self-reinforcing. Falling kidney function raises plasma oxalate, which increases deposition ^48^, and nephron loss itself drives further sclerosis through compensatory hyperfiltration ^49^. Consistently, the highest plasma oxalate quartile carried a 5- to 14-fold higher risk of ESKD in primary hyperoxaluria ^50^, and in a general CKD population each standard deviation increase in urinary oxalate was associated with a 9% higher risk of the composite kidney endpoint ^51^. Deposits also occur outside the kidney, including the retina and vasculature ^52,53^. Once clinically overt, reversibility is low: 55% of 108 cases required dialysis and none recovered renal function ^10^, 52% of a biopsy-confirmed cohort progressed to kidney failure within one month ^54^, and in the Mayo series 54% reached ESKD within a median of 36 months ^8^. The need for dialysis at diagnosis was the only multivariable predictor of a poor outcome ^47^.

Among patients who have not reached that endpoint the burden is still substantial: in 814 patients with EH followed for a mean of 5.0 years, the median stone event rate was 0.70 per year, rising to 1.24 in those with structural malabsorption, the group with the highest 5-year incidence of ESKD (17.8%) ^9^; bariatric surgery itself raises the long-term stone risk relative to the general population ^55^. The point in this chain most amenable to intervention lies upstream: oxalate restriction lowers urinary oxalate by about 21% ⁵⁶, and an oxalate-degrading enzyme lowers urinary oxalate in a randomised trial ⁵⁷, whereas the therapeutic window closes once kidney failure is reached. These prevalence estimates indicate which patient groups carry the highest burden of EH and therefore deserve priority when screening is considered.

### Estimated potential patient population in the United States

We applied these prevalences to the US populations in which both quantities are available: approximately 1.0 million people with CD,⁵⁸ 39,000 with CF,²¹ and 800,000 cumulative RYGB procedures.²⁶ This gives approximately 490,000 people with EH among those with CD, 21,000 with CF, and 440,000 after RYGB — about 950,000 in total. This should be read as an order of magnitude, not a count. Three sources of uncertainty preclude a firmer figure. First, every included study was a single-center series, most from tertiary referral clinics, so their prevalences need not apply to a national population. Second, the population denominators are themselves uncertain: published estimates of the number of people with CD in the United States differ (approximately 1.0 versus 1.3 million), and the cumulative RYGB figure rests on procedures performed within a reporting window. Third, the prevalences are heterogeneous summaries (I² > 90%) rather than single values, so the appropriate interval for application to a new population is the prediction interval, which places the total between approximately 240,000 and 1,650,000. A fourfold range cannot support service planning. The estimate also covers only these three groups; because CP, CeD, and SBS were not extrapolated, the total across all enteric causes would be higher.

For comparison, the only published estimate, derived from insurance claims rather than urinary oxalate, placed approximately 250,000 people with an enteric disorder or a history of enteric surgery as having kidney stones or CKD ^11^ — a count of those already harmed, not of those exposed. A similar reversal appears in the 814 patients of Tatton et al.^9^, where bariatric surgery accounted for 64% of the cohort and CeD for only 3%: kidney damage after bariatric surgery appears to run mainly through the oxalate pathway, whereas few patients with CeD reach the oxalate threshold. The disease groups were also clearly separated from healthy people, with no hyperoxaluria among the 45 controls of Salem et al.^17^ or the 38 of Scherer et al.^59^, and only 2 of 20 preoperative obese controls in Sinha et al.^42^, rising to 54.8% afterwards ^60,61^.

### Heterogeneity and subgroup analyses

All three pooled prevalences had I² above 90%, reflecting differences in threshold, age, resection proportion, stone history, diet, region, and year. Because most data come from hospital-based testing rather than population sampling, the pooled values are weighted averages across heterogeneous populations rather than universally applicable prevalences. Three factors account for most of the variation. Testing condition is the largest: crude prevalence was 26.1 percentage points higher when an oxalate load was given, and the pooled estimates differed by 34.9 points (74.0% vs 39.1%, Table 2), which is why the overall figure should not be applied to any single setting. Ileal resection status is next, at 17.1 points higher crude and 28.8 points higher pooled, with less heterogeneity in the non-resected subgroup (I² = 75.6% vs 94.0%, Table 3). Diagnostic threshold and population composition follow: after harmonising the threshold to 45 mg/24 h or its 0.5 mmol/24 h equivalent, six studies gave 42.1% (I² = 87.2%, Table 2), an interval overlapping that of all studies, so threshold alone does not explain the heterogeneity; age and stone selection could not be assessed comparably because the pediatric cohorts and the stone-selected cohorts differed from the comparators in geography or threshold as well. Two studies using a ratio or body-surface-area-corrected format gave 37.8% (17/45); excluding both left the pooled estimate essentially unchanged at 50.9% (39.1–62.6; 15 studies), and leave-one-out analysis kept the CD estimate between 45.7% and 51.8% (Figure S1).

### Limitations

#### Certainty of evidence

Using GRADE for single-proportion outcomes, certainty was very low for all three disease groups, downgraded for risk of bias (all cohorts were single-centre clinical or referral series, several recruited from stone-forming populations), for very serious inconsistency (I² 91.9%–96.7%, far beyond the 75% threshold at which our protocol specifies that pooling may not be appropriate), for indirectness (tertiary referral cohorts extrapolated nationally), and for imprecision (95% CI widths of 20.9, 19.7, and 18.6 percentage points). Funnel plots (Figures S2–S4) and Egger’s test showed no evidence of asymmetry (P = 0.105, 0.345, and 0.504), so publication bias was not a reason for downgrading, although the test has limited power in prevalence research. This is consistent with our prespecified approach: the pooled values are exploratory summaries and the narrative synthesis stratified by testing condition remains the primary result.

The review has further limitations. Included studies were referral cohorts rather than population samples, so selection bias is present. Andersson, Filipsson and Hultén (1978) were excluded from the pooled analysis because their cohort probably overlaps that of an included study from the same centre, and because all of their data were obtained after an oxalate load roughly twice that used elsewhere. Thresholds and collection formats were not uniform, some studies reported only a ratio not convertible to 24-hour excretion, and resection and steatorrhoea strata were available for only some studies. Our estimates rest on 24-hour urinary oxalate alone and omit other lithogenic factors such as urinary calcium, citrate, and volume; in bariatric studies, diet was generally uncontrolled and limb lengths differ between centres. Three studies recruited from stone-forming populations gave a higher crude prevalence (51.6%, 16/31) than the remaining cohorts (42.7%, 188/440), the direction expected from a selection effect, but the groups are not comparable and no test was performed. Finally, screening, extraction, and risk-of-bias assessment were performed by one investigator without duplicate review, and we did not search Chinese-language or grey literature.^62^

### Clinical implications

Assessment for EH should be considered in patients with CD, CF, or a history of bariatric surgery, particularly when steatorrhea, previous ileal resection, or recurrent kidney stones are present, although the certainty of the underlying estimates is very low (see Limitations). Steatorrhea, kidney stones, pancreatic atrophy, and a fecal acid steatocrit >31% indicate screening in patients with CP ^28^, and the first two extend to CD. Dietary conditions before collection must be standardized and reported: after a 500 mg oxalic acid or 100 g spinach load the detected proportion nearly doubles, so screening studies should use a sequential low-oxalate baseline followed by a high-oxalate load ^3,33^, and negative results from free-diet studies interpreted with caution. Because 24-hour collection is cumbersome, an oxalate-to-creatinine ratio can be used for initial screening, with positives confirmed by 24-hour collection.

The screening marker also needs qualification. The Kidney Health Initiative working group noted that urinary oxalate and calcium oxalate supersaturation can serve only as surrogate endpoints for symptomatic stone events, not as clinical endpoints ^63^, and in the 11 patients of Evan et al.^46^ only half had urinary oxalate above 45 mg/24 h. Screening and follow-up should therefore track stone events and kidney function, with urinary oxalate used to identify high-risk individuals rather than to grade severity.

## Conclusions

This review provides the first systematic synthesis of EH prevalence across CD, CF, and bariatric surgery, and shows that the measured prevalence depends heavily on how urine is collected: EH was present in approximately 39% of patients tested without an oxalate load but 65%–74% of those tested after one. Because most studies do not state this condition, the pooled estimates (49.3% for CD, 49.8% for CF, 54.7% after bariatric surgery; I² > 90%) are heterogeneous summaries rather than prevalences applicable to any single setting, and estimates reported elsewhere without a stated testing condition are not directly comparable. A history of ileal resection (48.8% vs 31.7%; OR 1.62, 95% CI 0.91–2.89) and the presence of steatorrhea (62.5% vs 29.3%; OR 9.88, 95% CI 3.81–25.61) were both associated with higher EH proportions and can be used as clinical stratification markers. Extrapolation to US population sizes suggests that several hundred thousand people with CD, CF, or a history of RYGB may have EH, but the estimate is too uncertain to serve as a count (range across assumptions, approximately 240,000 to 1,650,000); the size of this population has not been established. Because EH can progress to calcium oxalate stones, oxalate nephropathy, CKD, and even ESKD, establishing standardized urinary oxalate screening pathways in these disease populations has clear clinical value. Two principal gaps remain in the current evidence: the absence of population-based or prospective cohort prevalence data obtained under uniform diagnostic thresholds and dietary conditions; and the lack of agreement on the clinical endpoint for EH screening — whether a urinary oxalate threshold, stone events, or a decline in eGFR is used directly determines the cost-effectiveness of any screening program, and this question requires prospective study.

## Supporting information

FigureS1-S4_AppendixS1-S2

Data file and script

## Data Availability

All data produced in the present study are contained in the manuscript and its supplementary files: the study-level extraction workbook for all included studies, the full search strategies for PubMed/MEDLINE and Embase, the completed PRISMA 2020 checklist, and the analysis scripts. No individual patient data were used; all values are aggregate statistics extracted from published studies. The protocol is registered with PROSPERO (CRD420261364424).

## Acknowledgments

None.

## Author Contributions

Haifeng Liu and Qingshan Li conceived and designed the review; Xiaomin Xing performed the literature search, screening, data extraction, and statistical analysis; Huiqiong He independently verified a random 20% sample of the extracted data against the source publications; Xiaomin Xing drafted the manuscript. All authors read and approved the final manuscript.

## Use of Artificial Intelligence

Large language models (DeepSeek V4-Flash, 21 August 2026, and subsequent versions) were used to assist with this review. A large-language-model-assisted workflow was run independently of, and in parallel with, the principal investigator for title and abstract screening (agreement on definite include/exclude decisions, κ = 0.86). The models also assisted with full-text assessment, data extraction, statistical analysis, figure generation, and preparation of the manuscript text. All extracted values and all reported calculations were checked by the authors against the source publications and re-derived before inclusion. In accordance with ICMJE recommendations, the models are not listed as authors; the authors take full responsibility for the content.

## Ethics Statement

Not applicable; this systematic review used only previously published aggregate data.

## Supplementary material

**Figure S1.** Leave-one-out sensitivity analysis for Crohn’s disease (k = 17).

**Figures S2–S4.** Funnel plots for Crohn’s disease, cystic fibrosis, and bariatric surgery, with the corresponding Egger’s test results.

**Appendix S1.** Full search strategies for PubMed/MEDLINE and Embase.

**PRISMA 2020 checklist.** Completed checklist with the location of each item in the manuscript.

