## Supplementary material for "Prevalence of Enteric Hyperoxaluria in Crohn’s Disease, Cystic Fibrosis, and After Bariatric Surgery: A Systematic Review": FigureS1-S4_AppendixS1-S2

### Figure S1. Leave-one-out sensitivity analysis for Crohn's disease

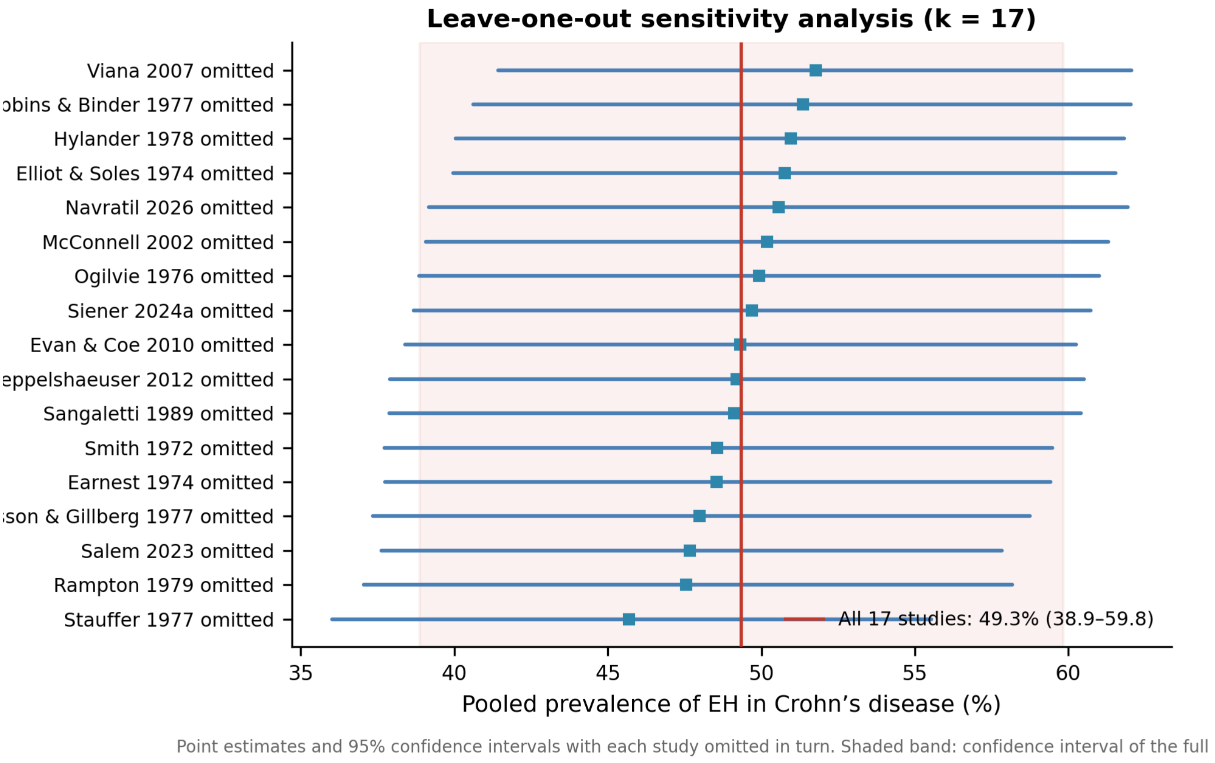


**Figure S1.** Leave-one-out sensitivity analysis for the pooled prevalence of enteric hyperoxaluria in Crohn's disease (k = 17). Each row shows the pooled estimate and 95% confidence interval obtained after omitting the study named on the y-axis. The vertical red line is the pooled estimate for all 17 studies (49.3%, 95% CI 38.9–59.8) and the shaded band its confidence interval. The pooled estimate ranged from 45.7% (omitting Stauffer 1977) to 51.8% (omitting Viana 2007); no single study changed the direction of the result.

**Figure S2. Funnel plot for Crohn's disease**

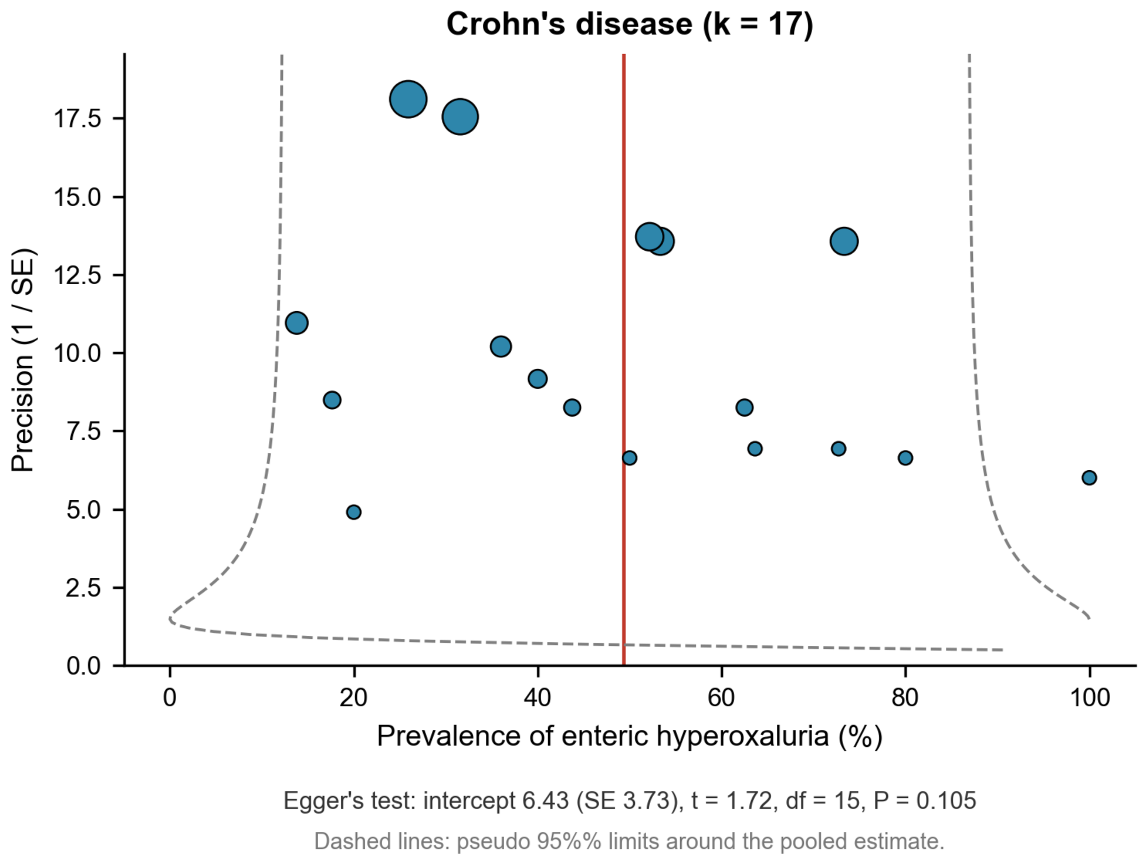


**Figure S2.** Funnel plot of study prevalence against precision for the 17 Crohn's disease studies. The vertical red line is the random-effects pooled estimate and the dashed lines the pseudo 95% limits around it; marker area is proportional to study sample size. Egger's test showed no statistical evidence of asymmetry (intercept 6.43, SE 3.73; P = 0.105).

**Figure S3. Funnel plot for cystic fibrosis**

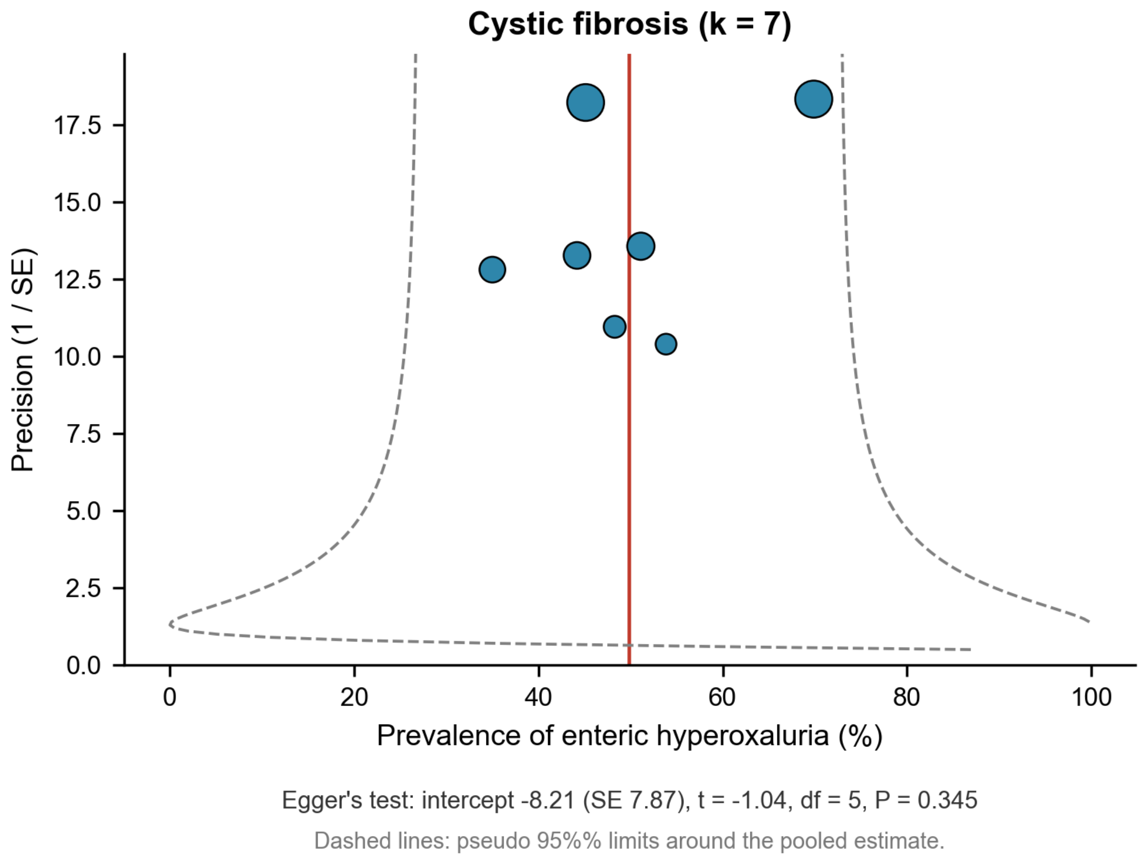


**Figure S3.** Funnel plot for the seven cystic fibrosis studies (pooled prevalence 49.8%, 95% CI 40.5–59.1). Conventions as in Figure S2. Egger's test showed no statistical evidence of asymmetry (intercept −8.21, SE 7.87; P = 0.345).

### Figure S4. Funnel plot for bariatric surgery

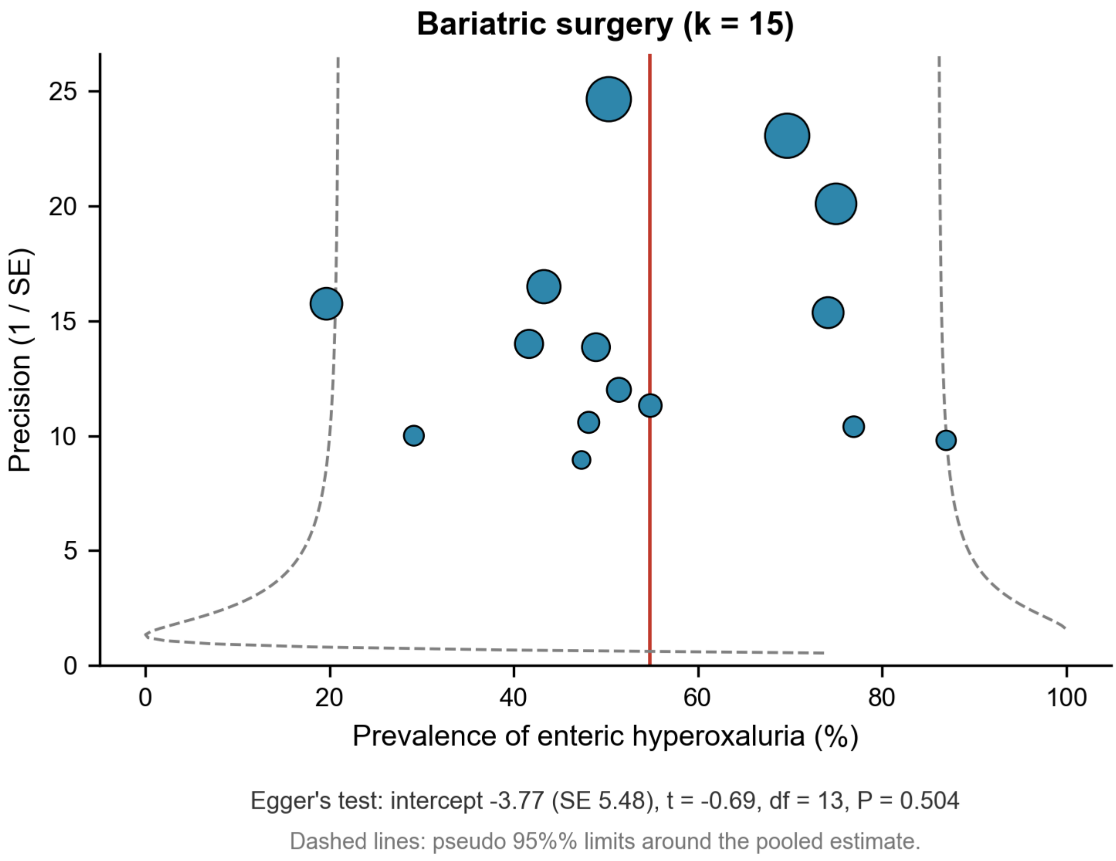


**Figure S4.** Funnel plot for the 15 studies after bariatric surgery. Conventions as in Figure S2. Egger's test showed no statistical evidence of asymmetry (intercept −3.77, SE 5.48; P = 0.504).

### Appendix S1. Search strategies

Databases were searched from inception to 19 August 2026 (PubMed) and 24 August 2026 (Embase), with no language restriction.

#### PubMed/MEDLINE

Concept blocks were combined as Block 1 AND (Block 2 OR Block 3 OR Block 4 OR Block 5 OR Block 6 OR Block 7).

##### Block 1 — oxalate

```
("hyperoxaluria"[MeSH] OR "hyperoxaluria"[tiab] OR "enteric
hyperoxaluria"[tiab] OR "absorptive hyperoxaluria"[tiab] OR
```

"oxaluria"[tiab] OR "urinary oxalate"[tiab] OR "oxalate excretion"[tiab] OR "oxalate absorption"[tiab])

**Blocks 2–7 — disease groups and fat malabsorption**

("Crohn disease"[MeSH] OR "Crohn's disease"[tiab] OR "Crohn disease"[tiab] OR "ileal resection"[tiab] OR "ileocolonic resection"[tiab] OR "short bowel syndrome"[MeSH] OR "short bowel"[tiab] OR "ileitis"[tiab]) OR ("pancreatitis, chronic"[MeSH] OR "chronic pancreatitis"[tiab] OR "exocrine pancreatic insufficiency"[tiab] OR "pancreatic steatorrhea"[tiab] OR "pancreatic malabsorption"[tiab]) OR ("cystic fibrosis"[MeSH] OR "cystic fibrosis"[tiab] OR "CFTR"[tiab] OR "mucoviscidosis"[tiab]) OR ("celiac disease"[MeSH] OR "coeliac disease"[tiab] OR "celiac disease"[tiab] OR "gluten enteropathy"[tiab] OR "nontropical sprue"[tiab]) OR ("bariatric surgery"[MeSH] OR "bariatric surgery"[tiab] OR "bariatric"[tiab] OR "gastric bypass"[MeSH] OR "gastric bypass"[tiab] OR "Roux-en-Y"[tiab] OR "roux en y"[tiab] OR "biliopancreatic diversion"[tiab] OR "duodenal switch"[tiab] OR "sleeve gastrectomy"[tiab] OR "jejunoileal bypass"[MeSH] OR "jejunoileal bypass"[tiab] OR "weight loss surgery"[tiab] OR "metabolic surgery"[tiab] OR "obesity surgery"[tiab] OR "malabsorptive surgery"[tiab]) OR "fat malabsorption"[tiab] OR "steatorrhea"[tiab] OR "steatorrhoea"[tiab] OR "malabsorption syndromes"[MeSH] OR "bile acid malabsorption"[tiab]

**Full strategy as executed**

(("hyperoxaluria"[MeSH] OR "hyperoxaluria"[tiab] OR "enteric hyperoxaluria"[tiab] OR "absorptive hyperoxaluria"[tiab] OR "oxaluria"[tiab] OR "urinary oxalate"[tiab] OR "oxalate excretion"[tiab] OR "oxalate absorption"[tiab]) AND (("Crohn disease"[MeSH] OR "Crohn's disease"[tiab] OR "Crohn disease"[tiab] OR "ileal resection"[tiab] OR "ileocolonic resection"[tiab] OR "short bowel syndrome"[MeSH] OR "short bowel"[tiab] OR "ileitis"[tiab]) OR ("pancreatitis, chronic"[MeSH] OR "chronic pancreatitis"[tiab] OR "exocrine pancreatic insufficiency"[tiab] OR "pancreatic steatorrhea"[tiab] OR "pancreatic malabsorption"[tiab]) OR ("cystic fibrosis"[MeSH] OR "cystic fibrosis"[tiab] OR "CFTR"[tiab] OR "mucoviscidosis"[tiab]) OR ("celiac disease"[MeSH] OR "coeliac disease"[tiab] OR "celiac disease"[tiab] OR "gluten enteropathy"[tiab] OR "nontropical sprue"[tiab]) OR ("bariatric surgery"[MeSH] OR "bariatric surgery"[tiab] OR "bariatric"[tiab] OR "gastric bypass"[MeSH] OR "gastric bypass"[tiab] OR "Roux-en-Y"[tiab] OR "roux en y"[tiab] OR "biliopancreatic diversion"[tiab] OR "duodenal switch"[tiab] OR "sleeve gastrectomy"[tiab] OR "jejunoileal bypass"[MeSH] OR "jejunoileal bypass"[tiab] OR "weight loss surgery"[tiab] OR "metabolic surgery"[tiab] OR "obesity surgery"[tiab] OR "malabsorptive surgery"[tiab]) OR "fat malabsorption"[tiab] OR "steatorrhea"[tiab] OR "steatorrhoea"[tiab] OR "malabsorption syndromes"[MeSH] OR "bile acid malabsorption"[tiab]))

Result: **365 records**.

**Embase (Command Line Search)**

('hyperoxaluria'/exp OR 'hyperoxaluria':ti,ab OR 'enteric hyperoxaluria':ti,ab OR 'absorptive hyperoxaluria':ti,ab OR 'oxaluria':ti,ab OR 'urinary oxalate':ti,ab OR 'oxalate excretion':ti,ab OR 'oxalate absorption':ti,ab) AND (('crohn disease'/exp OR 'crohn disease':ti,ab OR 'ileal resection':ti,ab OR 'ileocolonic resection':ti,ab OR 'short bowel syndrome'/exp OR 'short bowel':ti,ab OR 'ileitis':ti,ab) OR ('chronic pancreatitis'/exp OR 'chronic pancreatitis':ti,ab OR 'exocrine pancreatic insufficiency':ti,ab OR 'pancreatic steatorrhea':ti,ab OR 'pancreatic malabsorption':ti,ab) OR ('cystic

```
fibrosis'/exp OR 'cystic fibrosis':ti,ab OR 'cftr':ti,ab OR
'mucoviscidosis':ti,ab) OR ('celiac disease'/exp OR 'coeliac
disease':ti,ab OR 'celiac disease':ti,ab OR 'gluten enteropathy':ti,ab OR
'nontropical sprue':ti,ab) OR ('bariatric surgery'/exp OR 'bariatric
surgery':ti,ab OR 'bariatric':ti,ab OR 'gastric bypass'/exp OR 'gastric
bypass':ti,ab OR 'roux-en-y':ti,ab OR 'biliopancreatic diversion'/exp OR
'biliopancreatic diversion':ti,ab OR 'duodenal switch':ti,ab OR 'sleeve
gastrectomy'/exp OR 'sleeve gastrectomy':ti,ab OR 'jejunoileal bypass'/exp
OR 'jejunoileal bypass':ti,ab OR 'weight loss surgery':ti,ab OR 'metabolic
surgery':ti,ab OR 'obesity surgery':ti,ab OR 'malabsorptive surgery':ti,ab)
OR 'fat malabsorption':ti,ab OR 'steatorrhea':ti,ab OR
'steatorrhoea':ti,ab OR 'malabsorption syndrome'/exp OR 'bile acid
malabsorption':ti,ab)
```

Result: **820 records**.

**Yield of the concept blocks**

| Combination | Records |
|---|---|
| Oxalate AND Crohn's disease or short bowel | 104 |
| Oxalate AND cystic fibrosis | 35 |
| Oxalate AND chronic pancreatitis | 28 |
| Oxalate AND celiac disease | 34 |
| Oxalate AND bariatric surgery | 175 |
| Oxalate AND fat malabsorption only | 140 |

Records from the two databases were combined and de-duplicated in Rayyan: 1,185 identified, 312 duplicates removed, 873 screened. Reference lists of all retrieved articles were also hand-searched; this yielded 15 additional records, all assessed at full-text level and included among the studies reported.

### Appendix S2. PRISMA 2020 checklist

| Section / topic | Item | Checklist item | Location in manuscript |
|---|---|---|---|
| **TITLE** | | | |
| Title | 1 | Identify the report as a systematic review. | Title page: "…: A Systematic Review" |
| **ABSTRACT** | | | |
| Abstract | 2 | See the PRISMA 2020 for Abstracts checklist. | Abstract (structured: Introduction, Methods, Results, Conclusion) |
| **INTRODUCTION** | | | |
| Rationale | 3 | Describe the rationale for the review in the context of existing knowledge. | Introduction, paragraphs 1–4 |
| Objectives | 4 | Provide an explicit statement of the objective or question. | Introduction, final paragraph |
| **METHODS** | | | |
| Eligibility criteria | 5 | Specify the inclusion and exclusion criteria and how studies were grouped. | 2.1 |
| Information sources | 6 | Specify all databases, registers, websites, and | 2.1 (PubMed and Embase, to 24 August 2026); |

| Section / topic | Item | Checklist item | Location in manuscript |
|---|---|---|---|
| | | other sources, with the date last searched. | Appendix S1 |
| Search strategy | 7 | Present the full search strategies for all databases, including any filters and limits. | Appendix S1 |
| Selection process | 8 | Specify the methods used to decide whether a study met the criteria, including how many reviewers screened each record and whether they worked independently. | 2.1 (PI and an independently run large-language-model-assisted workflow; κ = 0.86) |
| Data collection process | 9 | Specify the methods used to collect data, including how many reviewers collected data and whether they worked independently. | 2.1, 2.2 |
| Data items | 10 | List and define all outcomes and other variables sought. | 2.2 |
| Risk of bias assessment | 11 | Specify the methods used to assess risk of bias, including the tool(s) used and how many reviewers assessed each study. | 2.3, 4.5 (GRADE for single-proportion outcomes; assessed by one investigator) |
| Effect measures | 12 | Specify for each outcome the effect measure(s) used. | 2.3 (prevalence; Freeman–Tukey double arcsine transformation) |
| Synthesis methods | 13a | Describe the processes used to decide which studies were eligible for each synthesis. | 2.3 ($k \geq 3$ for pooling; $k \leq 2$ crude only) |
| | 13b | Describe any methods used to tabulate or display results and synthesise them. | 2.3; Tables 1–8, Figures 1–4 |
| | 13c | Describe any methods used to explore possible causes of heterogeneity. | 2.3 (prespecified subgroups) |
| | 13d | Describe any sensitivity analyses conducted. | 2.3 (leave-one-out analysis; Figure S1) |
| | 13e | Describe any methods used to assess certainty in the body of evidence. | 2.3 (protocol item 4.8.2), 4.5 (GRADE) |
| | 13f | Describe any methods used to explore possible causes of heterogeneity. | 2.3; 4.4 |
| Reporting bias assessment | 14 | Describe any methods used to assess risk of bias due to missing results. | 4.5 (funnel plots and Egger's test for the three main analyses) |
| Certainty assessment | 15 | Describe any methods used to assess certainty in the body of evidence. | 2.3; 4.5 (GRADE, single-proportion framework) |
| **RESULTS** | | | |
| Study selection | 16a | Describe the results of the search and selection process, ideally with a flow diagram. | 3.1; Figure 1 |
| | 16b | Cite studies that appeared to meet the criteria but were excluded, and explain why. | 3.1; Table S2 (Andersson, Filipsson and Hultén 1978, excluded with reasons) |
| Study characteristics | 17 | Cite each included study and present its | Tables 1, 5, 6, 7; Table S1 |

| Section / topic | Item | Checklist item | Location in manuscript |
|---|---|---|---|
| | | characteristics. | |
| Risk of bias in studies | 18 | Present assessments of risk of bias for each included study. | 4.5 (GRADE domain summaries) |
| Results of individual studies | 19 | For all outcomes, present summary statistics for each study. | Tables 1, 4, 5, 6, 7, 8; Figures 2, 3, 4 |
| Results of syntheses | 20a | For each synthesis, briefly summarise the characteristics and risk of bias among contributing studies. | 3.2–3.5; 4.4 |
| | 20b | Present results of all statistical syntheses; if meta-analysis was done, present the summary estimate, its precision, and measures of heterogeneity. | 3.2, 3.3, 3.4 (pooled prevalence with 95% CI, $I^2$, $\tau^2$) |
| | 20c | Present results of investigations of possible causes of heterogeneity. | 3.2; Table 2; 4.4 |
| | 20d | Present results of any sensitivity analyses. | 4.4 (leave-one-out, 45.7%–51.8%); Figure S1; Table S2 |
| Reporting biases | 21 | Present assessments of risk of bias due to missing results. | 4.5 (Egger's test P = 0.105, 0.184, 0.504; funnel plots available on request — see note below) |
| Certainty of evidence | 22 | Present assessments of certainty in the body of evidence for each outcome assessed. | 4.5 (very low for all three disease groups, with reasons for downgrading) |
| **DISCUSSION** | | | |
| Discussion | 23a | Provide a general interpretation of the results and their implications. | 4.1–4.6 |
| | 23b | Discuss limitations of the evidence included in the review. | 4.5 |
| | 23c | Discuss limitations of the review processes. | 4.5 |
| | 23d | Discuss implications for practice, policy, and future research. | 4.6; 5 |
| **OTHER INFORMATION** | | | |
| Registration and protocol | 24a | Provide registration information, including the register name and registration number, or state that the review was not registered. | Title page; 2.1 (PROSPERO CRD420261364424) |
| | 24b | Indicate where the review protocol can be accessed, or state that it is not available. | 2.1; PROSPERO record |
| | 24c | Describe and explain any amendments to information provided at registration. | 2.3 (deviation: when $I^2 \geq$ 75%, pooled estimates are reported as exploratory summaries with narrative synthesis as the primary result) |

| Section / topic | Item | Checklist item | Location in manuscript |
|---|---|---|---|
| Support | 25 | Describe sources of financial or non-financial support and the role of the funders. | Funding statement |
| Competing interests | 26 | Declare any competing interests of review authors. | Disclosure |
| Availability of data, code, and other materials | 27 | Report which of the following are publicly available and where: template data collection forms, extracted data, analysis code, and any other materials. | Data sharing statement; Tables S1–S2; Appendix S1 |

### Notes

**PRISMA 2020 for Abstracts** (item 2). The abstract reports the review question, eligibility criteria, information sources, number of included studies, and a synthesis of results, in line with the PRISMA 2020 for Abstracts checklist.

**Reporting bias (items 14 and 21).** Funnel plots were generated for the three main analyses (Crohn's disease, k = 17; cystic fibrosis, k = 7; bariatric surgery, k = 15) and are available as supplementary material on request; Egger's test was not statistically significant for any of the three, so publication bias was not a reason for downgrading certainty. The test has limited power in prevalence research, and asymmetry in prevalence funnel plots cannot be attributed to publication bias alone.

**Risk of bias in individual studies (item 18).** Risk of bias was assessed with an adapted Newcastle–Ottawa scheme covering representativeness, sample size, exposure ascertainment, outcome assessment, and statistical reporting. Because the scheme was applied by a single investigator, the domain-level judgements reported in Section 4.5 under GRADE are presented in preference to study-level scores; the full scoring sheet is available from the corresponding author.

**Deviations from the protocol (item 24c).** The protocol prespecified meta-regression for subgroups with at least ten studies per covariate; this was not performed because no subgroup met that condition, and the protocol itself notes that meta-regression would not be undertaken in that case.
