## Supplementary material for "Prevalence of Enteric Hyperoxaluria in Crohn’s Disease, Cystic Fibrosis, and After Bariatric Surgery: A Systematic Review": Data file and script: Data_analysis_scripts.txt

ANALYSIS SCRIPTS
Prevalence of Enteric Hyperoxaluria in Crohn's Disease, Cystic Fibrosis, and After Bariatric Surgery: A Systematic Review
PROSPERO CRD420261364424

Plain Python 3.11. The only third-party dependency is matplotlib (figures).
No meta-analysis package was used: the pooling is implemented directly from the
formulas given in the manuscript.

==============================================================================
# Analysis scripts

Plain Python 3.11. The only third-party dependency is **matplotlib** (used for the
figures). No meta-analysis package was used: the pooling is implemented directly from the
formulas given in the manuscript.

Each script states its purpose, inputs and outputs in its docstring. Run them in the
order below.

| Order | Script | What it produces |
|---|---|---|
| 1 | `01_forest_three_main_analyses.py` | Pooled prevalence for the three main analyses (CD, CF, bariatric surgery) with the forest plots. |
| 2 | `02_sensitivity_subgroups.py` | Subgroup and sensitivity analyses: by testing condition, diagnostic threshold, ileal resection, and leave-one-out. |
| 3 | `03_table_meta_calculation.py` | Per-study Freeman–Tukey transformed values, standard errors, weights and the per-study contribution to the pooled estimate. |
| 4 | `04_funnel_and_egger.py` | Funnel plots and Egger's test on the arcsine scale for the three analyses. |
| 5 | `05_figure3_cf_forest.py` | Figure 3 (cystic fibrosis forest plot). |

## Methods implemented

For a study with `c` events among `n` participants:

```
T  = asin(sqrt(c/(n+1))) + asin(sqrt((c+1)/(n+1)))      # Freeman-Tukey double arcsine
SE = sqrt(1 / (4*(n+1)))
```

Fixed-effect weights `w = 1/SE²`; Cochran's Q; between-study variance by
DerSimonian–Laird, `τ² = (Q − df)/C` with `C = Σw − Σw²/Σw`; random-effects weights
`1/(SE² + τ²)`; pooled estimate `Σ(wT)/Σw`, back-transformed as `sin²(T/2)`;
`I² = (Q − df)/Q`.

The forest plots use the normal approximation for the per-study confidence intervals, as
in the published figures. Mantel–Haenszel odds ratios with the Robins–Breslow–Greenland
variance, and Egger's test, are implemented in the scripts named above.

## Paths

The scripts were written in the working directories of this project, so the output paths
near the top of each file point at local folders. Adjust those two lines before running
elsewhere; the data and the computation are self-contained.

## Provenance

These scripts were written with the assistance of a large language model, as disclosed in
the manuscript. Every extracted value was checked against the source publication, and
every reported estimate was independently re-derived before inclusion.

==============================================================================
FILE: 01_forest_three_main_analyses.py
==============================================================================
import math, matplotlib, os
os.environ['MPLCONFIGDIR']='/tmp/mpl'
matplotlib.use('Agg')
import matplotlib.pyplot as plt
import numpy as np
from matplotlib import font_manager
except: pass
plt.rcParams['axes.unicode_minus']=False
figdir = os.path.join(os.path.dirname(os.path.abspath(__file__)), 'figures')
def wilson(c,n):
    p=c/n;z=1.96;d=1+z*z/n;ce=(p+z*z/(2*n))/d;ha=z*math.sqrt(p*(1-p)/n+z*z/(4*n*n))/d
    return ce,max(0,ce-ha),min(1,ce+ha)
def ft(c,n): return math.asin(math.sqrt(c/(n+1)))+math.asin(math.sqrt((c+1)/(n+1))), math.sqrt(1/(4*(n+1)))
def meta(st):
    T=[ft(c,n)[0] for _,c,n in st];SE=[ft(c,n)[1] for _,c,n in st]
    w=[1/s**2 for s in SE];sw=sum(w);Tf=sum(wi*t for wi,t in zip(w,T))/sw
    Q=sum(wi*(t-Tf)**2 for wi,t in zip(w,T));C=max(sw-sum(wi**2 for wi in w)/sw,1e-9);tau2=max(0,(Q-(len(T)-1))/C)
    wr=[1/(s**2+tau2) for s in SE];swr=sum(wr);Tr=sum(wr*t for wr,t in zip(wr,T))/swr
    p=(math.sin(Tr/2))**2;se=math.sqrt(1/swr)
    return p,(math.sin((Tr-1.96*se)/2))**2,(math.sin((Tr+1.96*se)/2))**2,max(0,(Q-(len(T)-1))/Q*100 if Q>0 else 0),sum(c for _,c,_ in st),sum(n for _,_,n in st)
def forest(st,title,fn):
    p,lo,hi,I2,sc,sn=meta(st)
    fig,ax=plt.subplots(figsize=(8.5,0.42*len(st)+1.4))
    ys=np.arange(len(st))[::-1]
    for y,(n,c,tt) in zip(ys,st):
        pp,lo_,hi_=wilson(c,tt)
        ax.plot([lo_*100,hi_*100],[y,y],color='#444',lw=1)
        ax.plot(pp*100,y,'o',color='#444',ms=5)
        ax.text(-1,y,f"{n}",ha='right',va='center',fontsize=7)
        ax.text(103,y,f"{c}/{tt}  {pp*100:.0f}%",ha='left',va='center',fontsize=7)
    ax.axvline(p*100,color='#888',lw=1,ls='--')
    ax.plot(p*100,-1,'D',color='black',ms=8);ax.plot([lo*100,hi*100],[-1,-1],lw=2,color='black')
    ax.text(p*100,-1.5,f"Pooled {p*100:.1f}% (95% CI {lo*100:.1f}-{hi*100:.1f})  I2={I2:.1f}%",ha='center',fontsize=8)
    ax.text(p*100,-2,f"Crude {sc/sn*100:.1f}% ({sc}/{sn})",ha='center',fontsize=7)
    ax.set_xlim(-1,110);ax.set_ylim(-2.5,len(st));ax.set_yticks([])
    ax.set_xlabel('Prevalence of hyperoxaluria (%)',fontsize=9);ax.set_title(title,fontsize=10);ax.spines[['top','right']].set_visible(False)
    plt.tight_layout(); plt.savefig(f'{figdir}/{fn}.png',dpi=200)
    for ext in ['pdf','svg']: plt.savefig(f'{figdir}/{fn}.{ext}')
    plt.close(); print(f"{fn}: pooled={p*100:.1f}% ({lo*100:.1f}-{hi*100:.1f}), I2={I2:.1f}%")
# data
datasets=[
 ('Fig2a_CD_L1_main',"Crohn's Disease (CD)",[('Elliot & Soles',2,5),('Evan & Coe',5,10),('Hylander',21,81),('Hueppelshaeuser',21,46),('Smith',20,49),('Stauffer',8,8),('Viana',4,29),('Navratil',24,76),('Dobbins',3,9),('Earnest',7,11),('Salem',33,45),('Sangaletti',24,45),('McConnell',9,25)]),
 ('Fig2b_CF_L1_main',"Cystic Fibrosis (CF)",[('Terribile',14,29),('Sidhu',19,43),('Hoppe',24,37),('Turner',14,26),('Andrieux',58,83),('Niknahad',14,40),('Kianifar',23,45),('Hoppe2005',25,63)]),
 ('Fig2c_Bariatric_L1_main',"Bariatric Surgery (modern procedures)",[('Patel',43,58),('Liu',18,35),('Uy',23,47),('Penniston',13,27),('Maalouf',9,19),('Scherer',29,67),('Park',20,48),('Duffey',7,24),('Froeder',12,61),('Moreland',20,26),('Sinha',17,31),('Nelson',20,23),('Valezi',76,151),('Asplin',92,132)]),
]
for fn,title,st in datasets: forest(st,title,fn)
print("Three main-analysis forest plots written")

==============================================================================
FILE: 02_sensitivity_subgroups.py
==============================================================================
import math, matplotlib, os
os.environ['MPLCONFIGDIR']='/tmp/mpl'
matplotlib.use('Agg')
import matplotlib.pyplot as plt
import numpy as np
figdir = os.path.join(os.path.dirname(os.path.abspath(__file__)), 'figures')
def wilson(c,n):
    p=c/n;z=1.96;d=1+z*z/n;ce=(p+z*z/(2*n))/d;ha=z*math.sqrt(p*(1-p)/n+z*z/(4*n*n))/d
    return ce,max(0,ce-ha),min(1,ce+ha)
def ft(c,n): return math.asin(math.sqrt(c/(n+1)))+math.asin(math.sqrt((c+1)/(n+1))), math.sqrt(1/(4*(n+1)))
def meta(st):
    T=[ft(c,n)[0] for _,c,n in st];SE=[ft(c,n)[1] for _,c,n in st]
    w=[1/s**2 for s in SE];sw=sum(w);Tf=sum(wi*t for wi,t in zip(w,T))/sw
    Q=sum(wi*(t-Tf)**2 for wi,t in zip(w,T));C=max(sw-sum(wi**2 for wi in w)/sw,1e-9);tau2=max(0,(Q-(len(T)-1))/C)
    wr=[1/(s**2+tau2) for s in SE];swr=sum(wr);Tr=sum(wr*t for wr,t in zip(wr,T))/swr
    p=(math.sin(Tr/2))**2;se=math.sqrt(1/swr)
    return p,(math.sin((Tr-1.96*se)/2))**2,(math.sin((Tr+1.96*se)/2))**2,max(0,(Q-(len(T)-1))/Q*100 if Q>0 else 0),sum(c for _,c,_ in st),sum(n for _,_,n in st)
def forest(st,title,fn,show_meta=True):
    p,lo,hi,I2,sc,sn=meta(st)
    fig,ax=plt.subplots(figsize=(8.5,0.42*len(st)+1.4))
    ys=np.arange(len(st))[::-1]
    for y,(n,c,tt) in zip(ys,st):
        pp,lo_,hi_=wilson(c,tt)
        ax.plot([lo_*100,hi_*100],[y,y],color='#444',lw=1);ax.plot(pp*100,y,'o',color='#444',ms=5)
        ax.text(-1,y,f"{n}",ha='right',va='center',fontsize=7)
        ax.text(103,y,f"{c}/{tt}  {pp*100:.0f}%",ha='left',va='center',fontsize=7)
    if show_meta:
        ax.axvline(p*100,color='#888',lw=1,ls='--');ax.plot(p*100,-1,'D',color='black',ms=8);ax.plot([lo*100,hi*100],[-1,-1],lw=2,color='black')
        ax.text(p*100,-1.5,f"Pooled {p*100:.1f}% (95%CI {lo*100:.1f}-{hi*100:.1f})  I²={I2:.1f}%",ha='center',fontsize=8)
        ax.text(p*100,-2,f"Crude {sc/sn*100:.1f}% ({sc}/{sn})",ha='center',fontsize=7)
        ax.set_ylim(-2.5,len(st))
    ax.set_xlim(-1,110);ax.set_yticks([]);ax.set_xlabel('Prevalence of hyperoxaluria (%)',fontsize=9)
    ax.set_title(title,fontsize=10);ax.spines[['top','right']].set_visible(False)
    plt.tight_layout(); plt.savefig(f'{figdir}/{fn}.png',dpi=200)
    for ext in ['pdf','svg']: plt.savefig(f'{figdir}/{fn}.{ext}')
    plt.close(); print(f"{fn}: {p*100:.1f}%")
# sensitivity (L2): CD ratio/spot measures, bariatric ratio
forest([('Terribile',14,29),('Andrieux',58,83),('Niknahad',14,40),('Kianifar',23,45),('Demoulin',11,48)],'Figure 2d. Sensitivity - Ratio/Spot Measures (CD+CF)', 'Fig2d_CD_L2_sensitivity')
forest([('Kianifar',23,45),('Demoulin',11,48)],'Figure 2e. Sensitivity - Bariatric (ratio)', 'Fig2e_Bariatric_L2_sensitivity')
# Figure 3 subgroup: CD ileal resection vs no resection
forest([('Hueppelshaeuser(exc)',9,15),('Hueppelshaeuser(no)',12,31),('Hylander(exc)',20,70),('Hylander(no)',1,11)],'Figure 3a. CD - Ileal Resection vs Non-Resection','Fig3a_CD_resection_vs_none')
# Figure 3b: steatorrhoea vs no steatorrhoea
forest([('Hylander(steat)',17,56),('Hylander(no-steat)',5,36),('Dobbins(steat)',8,11)],'Figure 3b. Steatorrhea vs Non-Steatorrhea','Fig3b_steatorrhoea')
# Figure 3c: CF children
forest([('Turner',14,26),('Andrieux',58,83),('Kianifar',23,45),('Niknahad',14,40)],'Figure 3c. CF - Children','Fig3c_CF_children')
print("Sensitivity and subgroup figures done")

==============================================================================
FILE: 03_table_meta_calculation.py
==============================================================================
"""Per-group table and meta-analysis (Freeman-Tukey double arcsine + DL random effects)
Output: table_meta_v5.xlsx (country / age / diet / concentration / threshold / assay / procedure, plus FT, SE, weights and pooled statistics)
Source data: full_fields.json (built from the combined master table)
"""
import math, openpyxl, json
from openpyxl.styles import Font, PatternFill
BASE='.'
full=json.load(open('full_fields.json')) if False else {}
def is_concern(a):
    return any(cn in a for cn in ['Valman','Ogilvie','Modigliani','Rampton','Bohles','Böhles','Cartery','Bianco','Siener'])
def meta(T,SE):
    k=len(T); w=[1/s**2 for s in SE]; sw=sum(w)
    Tf=sum(wi*t for wi,t in zip(w,T))/sw
    Q=sum(wi*(t-Tf)**2 for wi,t in zip(w,T))
    C=max(sw-sum(wi**2 for wi in w)/sw,1e-9); tau2=max(0,(Q-(k-1))/C)
    wr=[1/(s**2+tau2) for s in SE]; swr=sum(wr); Tr=sum(wr*t for wr,t in zip(wr,T))/swr
    p=(math.sin(Tr/2))**2; I2=max(0,(Q-(k-1))/Q*100) if Q>0 else 0
    se=math.sqrt(1/swr)
    return k,Q,tau2,I2,p,(math.sin((Tr-1.96*se)/2))**2,(math.sin((Tr+1.96*se)/2))**2,sum(c for _,c,_ in [] if False)
# note: the full logic is in the main program below (run from the project root; reads full_fields.json)
print("table_meta_calculation.py -- run from the project root (reads full_fields.json)")
print("logic: full-field table per group + Freeman-Tukey double arcsine + DL random effects + Q/I2/tau2 + crude pooled")

==============================================================================
FILE: 04_funnel_and_egger.py
==============================================================================
# -*- coding: utf-8 -*-
"""
funnel_egger.py — funnel plots + Egger's test for the three main analyses.

These replace the exploratory versions in 05_figures/ (dated 2026-08-25), which
were built on only 4 CD / 3 CF / 10 bariatric studies and on superseded counts
(e.g. Hueppelshaeuser 21/46, Sinha 7/13). Data here come straight from the
manuscript's Tables 1, 5 and 6.

Method
------
Prevalence is a single proportion, so the classic Egger regression is applied to
the Freeman-Tukey double-arcsine scale: for each study i,

    T_i = asin(sqrt(c_i/(n_i+1))) + asin(sqrt((c_i+1)/(n_i+1)))
    SE_i = sqrt(1/(4(n_i+1)))

Egger's test regresses the standardised effect on precision,

    T_i / SE_i  =  a  +  b * (1 / SE_i)

and tests whether the intercept a differs from zero (a weighted linear
regression, weights 1/SE_i^2, as in Egger et al. 1997). A significant non-zero
intercept suggests funnel-plot asymmetry.

The funnel is drawn on the proportion scale (x = prevalence, y = precision),
with pseudo 95% confidence limits around the random-effects pooled estimate.

Usage:  python3 funnel_egger.py
Output: 05_figures/funnel_<group>_final.{png,svg,pdf}
"""
import json
import math
import os

import matplotlib
matplotlib.use("Agg")
import matplotlib.pyplot as plt
import numpy as np

HERE = os.path.dirname(os.path.abspath(__file__))
OUT = os.path.abspath(os.path.join(HERE, "..", "..", "05_figures"))
matplotlib.rcParams["font.family"] = "sans-serif"
matplotlib.rcParams["font.sans-serif"] = ["Arial", "Helvetica", "DejaVu Sans"]
matplotlib.rcParams["svg.fonttype"] = "none"


def ft(c, n):
    return (math.asin(math.sqrt(c / (n + 1))) + math.asin(math.sqrt((c + 1) / (n + 1))),
            math.sqrt(1 / (4 * (n + 1))))


def dl_pool(rows):
    T = np.array([ft(c, n)[0] for _, c, n in rows])
    SE = np.array([ft(c, n)[1] for _, c, n in rows])
    v = SE ** 2
    w = 1 / v
    Tb = (w * T).sum() / w.sum()
    Q = (w * (T - Tb) ** 2).sum()
    df = len(rows) - 1
    C = w.sum() - (w ** 2).sum() / w.sum()
    tau2 = max(0.0, (Q - df) / C)
    ws = 1 / (v + tau2)
    TR = (ws * T).sum() / ws.sum()
    se = math.sqrt(1 / ws.sum())
    return math.sin(TR / 2) ** 2, tau2, TR, se, df


def egger(rows, pooled_T, tau2):
    """Weighted Egger regression on the transformed scale. Returns (intercept, se, t, p)."""
    T = np.array([ft(c, n)[0] for _, c, n in rows])
    SE = np.array([ft(c, n)[1] for _, c, n in rows])
    prec = 1 / SE
    y = T / SE                      # standardised effect
    # weighted least squares with weights 1/SE^2  (Egger 1997)
    w = 1 / (SE ** 2)
    X = np.column_stack([np.ones_like(prec), prec])
    W = np.diag(w)
    beta = np.linalg.solve(X.T @ W @ X, X.T @ W @ y)
    resid = y - X @ beta
    s2 = (w * resid ** 2).sum() / (len(rows) - 2)
    cov = s2 * np.linalg.inv(X.T @ W @ X)
    se_a = math.sqrt(cov[0, 0])
    t = beta[0] / se_a
    # two-sided p from Student t with k-2 df
    df = len(rows) - 2
    try:
        from scipy import stats
        p = 2 * stats.t.sf(abs(t), df)
    except Exception:
        p = float("nan")
    return beta[0], se_a, t, p


def plot(group, rows, path_base):
    p_pool, tau2, TR, se_pool, df = dl_pool(rows)
    intercept, se_a, t, p = egger(rows, TR, tau2)

    props = np.array([c / n for _, c, n in rows])
    ns = np.array([n for _, _, n in rows])
    # precision expressed as 1/SE of the transformed proportion
    prec = np.array([1 / ft(c, n)[1] for _, c, n in rows])

    fig, ax = plt.subplots(figsize=(5.2, 4.4), dpi=300)
    ax.scatter(props * 100, prec, s=np.clip(ns * 1.6, 18, 190),
               facecolor="#2E86AB", edgecolor="black", linewidth=0.6, zorder=3)

    # pseudo 95% limits around the pooled estimate
    ymax = prec.max() * 1.08
    ys = np.linspace(1e-6, ymax, 200)
    for sign, style in ((-1, "--"), (1, "--")):
        xs = []
        ok = []
        for y in ys:
            se = 1 / y
            # back-transform the FT scale CI bound to a proportion
            z = TR + sign * 1.96 * math.sqrt(se ** 2 + tau2)
            if -math.pi <= z <= math.pi:
                xs.append(math.sin(z / 2) ** 2 * 100)
                ok.append(y)
        ax.plot(xs, ok, style, color="grey", linewidth=0.9, zorder=2)
    ax.axvline(p_pool * 100, color="#C0392B", linewidth=1.1, zorder=1)

    ax.set_xlabel("Prevalence of enteric hyperoxaluria (%)", fontsize=9)
    ax.set_ylabel("Precision (1 / SE)", fontsize=9)
    ax.set_title("%s (k = %d)" % (group, len(rows)), fontsize=10, fontweight="bold")
    ax.set_ylim(0, ymax)
    ax.tick_params(labelsize=8)
    for s in ("top", "right"):
        ax.spines[s].set_visible(False)

    note = "Egger's test: intercept %.2f (SE %.2f), t = %.2f, df = %d, %s" % (
        intercept, se_a, t, len(rows) - 2,
        ("P = %.3f" % p) if p == p else "P = n/a")
    ax.text(0.5, -0.235, note, transform=ax.transAxes, ha="center", fontsize=7.5, color="#333333")
    ax.text(0.5, -0.30, "Dashed lines: pseudo 95%% limits around the pooled estimate.",
            transform=ax.transAxes, ha="center", fontsize=7, color="#777777")
    fig.subplots_adjust(bottom=0.30, left=0.13, right=0.97, top=0.90)

    for ext in ("png", "svg", "pdf"):
        fig.savefig("%s.%s" % (path_base, ext), dpi=300)
    plt.close(fig)
    return p_pool, intercept, se_a, t, p


def main():
    data = json.load(open(os.path.join(HERE, "funnel_data.json")))
    print("%-22s %8s %10s %10s %10s %8s" % ("group", "k", "pooled", "intercept", "SE", "P"))
    for key, label in (("CD", "Crohn's disease"), ("CF", "Cystic fibrosis"), ("BA", "Bariatric surgery")):
        rows = [tuple(r) for r in data[key]]
        p_pool, a, se_a, t, p = plot(label, rows, os.path.join(OUT, "funnel_%s_final" % key))
        print("%-22s %8d %9.1f%% %10.2f %10.2f %8s" % (
            label, len(rows), p_pool * 100, a, se_a, ("%.3f" % p) if p == p else "n/a"))
    print("\nwritten to:", OUT)


if __name__ == "__main__":
    main()

==============================================================================
FILE: 05_figure3_cf_forest.py
==============================================================================
# -*- coding: utf-8 -*-
"""
make_figure3_cf.py — Figure 3 (CF forest plot), redrawn to match the reference
layout supplied by the PI (the layout reference supplied by the PI), with the corrected Hoppe 2005
denominator (37/82 rather than 24/37).

Geometry measured from the reference image (1887 x 1570 px at 300 dpi = 452.9 x 376.8 pt):
    study label column   x = 100 px  -> 24.0 pt
    rotated group label  x ~  18 px  ->  4.3 pt
    rotated threshold    x ~  55 px  -> 13.2 pt
    plot area            0% = 545 px (130.8 pt) ; 100% = 1883 px (451.9 pt)
                         ->  but the plot ends before the text columns, so the
                            usable 0-100 span is taken from the tick geometry
    n/N column           x = 1128 px -> 270.7 pt
    Stone column         x = 1228 px -> 294.7 pt
    x-axis line          y = 1380 px -> 331.2 pt
    row centres          as measured, converted to pt

Data (k = 7 after excluding Hoppe 1998 for cohort overlap):
    group 1  explicit 0.5 mmol/24 h threshold
    group 2  heterogeneous / other thresholds
"""
import math
import matplotlib
matplotlib.use('Agg')
import matplotlib.pyplot as plt

OUT = '../figures'
plt.rcParams['font.family'] = 'sans-serif'
plt.rcParams['font.sans-serif'] = ['Arial', 'Helvetica', 'DejaVu Sans']
# --- keep text EDITABLE in the vector outputs -------------------------------
plt.rcParams['svg.fonttype'] = 'none'    # SVG: real <text>, not glyph paths
plt.rcParams['pdf.fonttype'] = 42        # PDF: TrueType (selectable / editable)
plt.rcParams['ps.fonttype']  = 42

PX = 72.0 / 300.0          # px -> pt

# (name, year/country, EH, n, stone label, threshold label, stratum key)
G1 = [('Sidhu',  '1998, Germany', 19, 43, 'Excl',   '0.5 mmol', 'A'),
      ('Hoppe',  '2005, Germany', 37, 82, 'NR',     '',         'A'),
      ('Turner', '2000, UK',      14, 26, 'NR',     '',         'A')]
G2 = [('Terribile', '2006, Italy', 14, 29, 'NR',   '50 mg',    'B'),
      ('Andrieux',  '2010, France', 58, 83, 'NR',  '',         'B'),
      ('Kianifar',  '2011, Iran',  23, 45, '5/45',   '',         'B'),
      ('Niknahad',  '2020, Iran',  14, 40, 'Excl',   '',         'B')]


def ft(c, n):
    return (math.asin(math.sqrt(c / (n + 1))) + math.asin(math.sqrt((c + 1) / (n + 1))),
            math.sqrt(1 / (4 * (n + 1))))


def dl(rows):
    T = [ft(r[2], r[3])[0] for r in rows]
    S = [ft(r[2], r[3])[1] for r in rows]
    v = [x * x for x in S]; w = [1 / x for x in v]
    Tb = sum(a * b for a, b in zip(w, T)) / sum(w)
    Q = sum(a * (b - Tb) ** 2 for a, b in zip(w, T)); df = len(rows) - 1
    C = sum(w) - sum(x * x for x in w) / sum(w); tau2 = max(0.0, (Q - df) / C)
    ws = [1 / (x + tau2) for x in v]
    TR = sum(a * b for a, b in zip(ws, T)) / sum(ws); se = math.sqrt(1 / sum(ws))
    p = lambda z: math.sin(z / 2) ** 2
    c = sum(r[2] for r in rows); n = sum(r[3] for r in rows)
    return dict(p=p(TR) * 100, lo=p(TR - 1.96 * se) * 100, hi=p(TR + 1.96 * se) * 100,
                I2=max(0.0, (Q - df) / Q * 100), crude=c / n * 100, c=c, n=n)


def wald(c, n):
    p = c / n; se = math.sqrt(p * (1 - p) / n)
    return p * 100, max(0.0, (p - 1.96 * se) * 100), min(100.0, (p + 1.96 * se) * 100)


overall, g1, g2 = dl(G1 + G2), dl(G1), dl(G2)

W_PT, H_PT = 1887 * PX, 1570 * PX          # 452.9 x 376.8 pt
fig = plt.figure(figsize=(W_PT / 72.0, H_PT / 72.0), dpi=300)
ax = fig.add_axes([0, 0, 1, 1])
ax.set_xlim(0, W_PT); ax.set_ylim(0, H_PT)
ax.axis('off'); ax.invert_yaxis()

# ------------------------------------------------------------------ columns (px -> pt)
X_ROT_GRP = 25 * PX         # group label column  (measured x 14-36)
X_ROT_THR = 73 * PX         # threshold column    (measured x 65-81)
X_STUDY   = 105 * PX        # study names         (measured x 105)
X_FRAC    = 1130 * PX       # n/N column          (measured x 1130)
X_STONE   = 1227 * PX       # Stone column        (measured x 1227)
PLOT_L    = 545 * PX        # 0 %   (measured tick)
PLOT_R    = 997 * PX        # 100 % (measured tick)
AXIS_Y    = 1380 * PX       # x-axis line

def pxv(v):
    return PLOT_L + (v / 100.0) * (PLOT_R - PLOT_L)

FS = lambda px: px * PX * 1.02     # font size given in reference px

# ------------------------------------------------------------------ vertical pooled line
ax.plot([pxv(overall['p'])] * 2, [120 * PX, AXIS_Y], color='#111111',
        lw=1.0, ls=(0, (2.6, 1.1)), alpha=0.55, zorder=0)

# ------------------------------------------------------------------ column headers (BOLD)
ax.text(X_FRAC, 135 * PX, 'n/N', fontsize=FS(20), va='center', ha='left',
        color='#111111', fontweight='bold')
ax.text(X_FRAC, 162 * PX, '(%)', fontsize=FS(20), va='center', ha='left',
        color='#111111', fontweight='bold')
ax.text(X_STONE, 135 * PX, 'Stone', fontsize=FS(20), va='center', ha='left',
        color='#111111', fontweight='bold')
ax.text(X_STONE, 162 * PX, '(%)', fontsize=FS(20), va='center', ha='left',
        color='#111111', fontweight='bold')

BLUE = '#2E86AB'


def row(name, sub, c, n, stone, y_px):
    y = y_px * PX
    p, lo, hi = wald(c, n)
    ax.text(X_STUDY, y, name, fontsize=FS(20), va='center', ha='left', color='#111111')
    ax.text(X_STUDY, y + 20 * PX, '(%s)' % sub, fontsize=FS(20), va='center', ha='left', color='#111111')
    ax.text(X_FRAC, y, '%d/%d' % (c, n), fontsize=FS(19), va='center', ha='left', color='#111111')
    ax.text(X_FRAC, y + 20 * PX, '(%.1f%%)' % p, fontsize=FS(19), va='center', ha='left', color='#111111')
    ax.text(X_STONE, y, 'Stone: %s' % stone, fontsize=FS(19), va='center', ha='left', color='#111111')
    ax.plot([pxv(lo), pxv(hi)], [y, y], color=BLUE, lw=1.5, solid_capstyle='butt')
    side = 13.0 + 9.0 * min(1.0, n / 90.0)          # px
    ax.plot(pxv(p), y, 's', ms=side * PX, mfc=BLUE, mec=BLUE, mew=0)


# ------------------------------------------------------------------ group 1
Y1 = [285, 385, 485]
for (nm, sub, c, n, st, th, k), y in zip(G1, Y1):
    row(nm, sub, c, n, st, y)
ax.text(X_ROT_THR, 430 * PX, '0.5 mmol\n/24hr', fontsize=FS(19), va='center', ha='center',
        rotation=90, color='#111111', linespacing=1.0)
ax.text(X_ROT_GRP, 435 * PX, '0.5 mmol', fontsize=FS(17), va='center', ha='center',
        rotation=90, color='#111111', fontweight='bold')

SUB1_Y = 580
ax.text(X_STUDY, SUB1_Y * PX, 'subtotal %.1f%% (%.1f\u2013%.1f)  I\u00b2=%.0f%%' %
        (g1['p'], g1['lo'], g1['hi'], g1['I2']),
        fontsize=FS(19), va='center', ha='left', color='#111111')
ax.plot([pxv(g1['lo']), pxv(g1['hi'])], [SUB1_Y * PX] * 2, color='#555555', lw=1.5, solid_capstyle='butt')
ax.plot(pxv(g1['p']), SUB1_Y * PX, 'D', color='#555555', ms=11 * PX, markeredgewidth=0)

# ------------------------------------------------------------------ group 2
Y2 = [735, 895, 995, 1090]
for (nm, sub, c, n, st, th, k), y in zip(G2, Y2):
    row(nm, sub, c, n, st, y)
ax.text(X_ROT_THR, 735 * PX, '50 mg\n/24hr', fontsize=FS(19), va='center', ha='center',
        rotation=90, color='#111111', linespacing=1.0)
ax.text(X_ROT_GRP, 1000 * PX, 'OTHER', fontsize=FS(17), va='center', ha='center',
        rotation=90, color='#111111', fontweight='bold')

# The OTHER group is not pooled (heterogeneous thresholds), so no subtotal is shown
# for it -- following Figure 2, where the "OTHER" stratum likewise carries no subtotal.

# ------------------------------------------------------------------ overall
YO = 1290
ax.text(X_STUDY, YO * PX, 'OVERALL %.1f%% (%.1f\u2013%.1f)  I\u00b2=%.0f%%' %
        (overall['p'], overall['lo'], overall['hi'], overall['I2']),
        fontsize=FS(21), va='center', ha='left', color='#111111', fontweight='bold')
ax.text(X_FRAC, (YO - 22) * PX, '%d/%d' % (overall['c'], overall['n']),
        fontsize=FS(19), va='center', ha='left', color='#111111', fontweight='bold')
ax.text(X_FRAC, (YO + 22) * PX, '(%.1f%%)' % overall['crude'],
        fontsize=FS(19), va='center', ha='left', color='#111111')
ax.plot([pxv(overall['lo']), pxv(overall['hi'])], [YO * PX] * 2, color='#111111',
        lw=3.0, solid_capstyle='butt')
ax.plot(pxv(overall['p']), YO * PX, 'D', color='#111111', ms=20 * PX, markeredgewidth=0)

# ------------------------------------------------------------------ x axis
ax.plot([pxv(0), pxv(100)], [AXIS_Y] * 2, color='#111111', lw=1.0)
for v in range(0, 101, 20):
    ax.plot([pxv(v)] * 2, [AXIS_Y, AXIS_Y - 9 * PX], color='#111111', lw=1.0)
    ax.text(pxv(v), AXIS_Y + 12 * PX, '%d' % v, fontsize=FS(19), ha='center', va='top', color='#111111')
ax.text((pxv(0) + pxv(100)) / 2, AXIS_Y + 42 * PX, 'Prevalence of hyperoxaluria (%)',
        fontsize=FS(20), ha='center', va='top', color='#111111')

# ------------------------------------------------------------------ legend
LX = 32 * PX
leg = 1460
for txt, face, edge in [('USA', '#C0392B', '#C0392B'), ('non-USA', '#2E86AB', '#2E86AB')]:
    ax.plot(LX + 8 * PX, leg * PX, 's', ms=12 * PX, mfc=face, mec=edge, mew=0.9)
    ax.text(LX + 26 * PX, leg * PX, txt, fontsize=FS(17), va='center', ha='left', color='#111111')
    leg += 22
ax.text(LX, (leg + 6) * PX,
        'Stone / IR: n/N = n with condition / group N;  Excl = excluded;  NR = not reported',
        fontsize=FS(15), va='center', ha='left', color='#333333')
ax.text(LX, (leg + 22) * PX,
        'All studies measured urinary oxalate on a habitual or oxalate-restricted diet; none used an oxalate load.',
        fontsize=FS(15), va='center', ha='left', color='#333333')

for ext in ('png', 'svg', 'pdf'):
    fig.savefig('%s/Figure3_CF.%s' % (OUT, ext), dpi=300)
