## Supplementary material for "Prevalence of Enteric Hyperoxaluria in Crohn’s Disease, Cystic Fibrosis, and After Bariatric Surgery: A Systematic Review": Data file and script: Data_readme.docx

**Data availability package**

PROSPERO CRD420261364424

### **What is in this package**

| **File** | **Contents** |
| --- | --- |
| S1_Extracted_study_data.xlsx | Study-level extraction for all three pooled analyses (CD, CF, bariatric surgery). Three worksheets. |
| Data_analysis_scripts.txt | The scripts used to compute the pooled estimates, the subgroup analyses, Egger's tests and the figures. |
| S1_README.md | This file. |

The full search strategies (**Appendix S1**) and the completed **PRISMA 2020 checklist**

(**Appendix S2**) are provided in the supplementary PDF that accompanies this manuscript.

### **S1 — the extracted data workbook**

#### **Columns**

Each worksheet begins with study identification (First author, Year, Country,

Centre and department, Design, Patient source), then the inclusion and exclusion

criteria as reported in the source paper, then the quantitative fields that entered the

analysis (Total patients, the number with enteric hyperoxaluria, the diagnostic

threshold, the diet before urine collection), then the outcome fields.

Two columns were added for this deposit:

- **Included in pooled analysis** — Yes or No.

- **Reason if not pooled** — for every study marked No.

Rows marked No are shaded. They are present because the workbook is the complete

full-text extraction record, not only the studies that were pooled. Without them the

worksheet totals would not match the manuscript.

#### **Reconciling the workbook totals with the manuscript**

| **Analysis** | **Included studies (k)** | **Patients** | **Manuscript** |
| --- | --- | --- | --- |
| Crohn's disease | 17 | 471 | k = 17, 471 patients |
| Cystic fibrosis | 7 | 348 | k = 7, 348 patients |
| Bariatric surgery | 15 | 849 | k = 15, 849 patients |

#### **Studies recorded but not pooled**

**Andersson, Filipsson and Hultén 1978** (*Scand J Gastroenterol* 13(4):465–469).

63 patients with Crohn's disease, all studied after an oxalate load of 800–900 mg/day,

roughly twice that used by any other included study, and probable cohort overlap with the

included Andersson and Gillberg 1977 study from the same centre. It is listed as a row

with Included in pooled analysis = No; no count from it was used in any analysis.

**Hoppe 1998** (*Pediatr Nephrol* 12:275–279). Cohort overlap with Hoppe 2005, which

states that 21 of its 37 patients had been followed since 1993 and had appeared in

earlier work.

#### **Note on Hoppe 2005**

Hoppe et al. 2005 (*Am J Kidney Dis* 46:440–445) screened 82 patients with cystic

fibrosis who were under care at one centre and found 37 who were persistently

hyperoxaluric (urinary oxalate >0.5 mmol/1.73 m²/24 h over the preceding 12 months).

Those 37 were studied in detail, and 24 of them were still hyperoxaluric at the time of

study.

The workbook uses **37/82** as the prevalence contribution, because 82 is the unselected

population denominator. The figure 24/37 is a follow-up proportion within a subgroup

selected for the outcome and is not a prevalence. Using 24/37 would have raised the

pooled cystic fibrosis estimate from 49.8% to 52.6%.

One consequence should be noted when reading the CF sheet: the 12-month criterion for

persistent hyperoxaluria is stricter than the single 24-hour measurement used by the

other six CF studies, so the CF estimate is the more conservative of the two readings.

### **S4 — analysis scripts**

The scripts are plain Python 3.11. Each states its inputs and outputs at the top.

S4_Analysis_scripts/README_scripts.md describes the order in which to run them.

Two things should be said plainly about provenance:

- The statistical analysis is a direct implementation of the methods described in the

manuscript: the Freeman–Tukey double arcsine transformation, a DerSimonian–Laird

random-effects model, Cochran's Q, I² and τ², Mantel–Haenszel odds ratios with

Robins–Breslow–Greenland variance, and Egger's test on the arcsine scale. No

third-party meta-analysis package was used.

- The scripts were written with the assistance of a large language model, as disclosed in

the manuscript. Every extracted value was checked against the source publication and

every reported estimate was re-derived before inclusion.

### **Citing this package**

If a DOI has been minted for this deposit, cite that DOI. Otherwise cite the manuscript.

### **Contact**

Corresponding author: Qingshan Li, Wuhan Kangfude Biotechnology Co., Ltd.,

Jiangxia District, Wuhan, Hubei Province 430000, China.
